# A system biology approach identifies candidate drugs to reduce mortality in severely ill COVID-19 patients

**DOI:** 10.1101/2021.09.14.21262309

**Authors:** Vinicius M. Fava, Mathieu Bourgey, Pubudu M. Nawarathna, Marianna Orlova, Pauline Cassart, Donald C. Vinh, Matthew Pellan Cheng, Guillaume Bourque, Erwin Schurr, David Langlais

## Abstract

Despite the availability of highly efficacious vaccines, Coronavirus Disease 2019 (COVID-19) caused by severe acute respiratory syndrome-related coronavirus-2 (SARS-CoV-2) lacks effective drug treatment which results in a high rate of mortality. To address this therapeutic shortcoming, we applied a system biology approach to the study of patients hospitalized with severe COVID. We show that, at the time of hospital admission, patients who were equivalent on the clinical ordinal scale displayed significant differential monocyte epigenetic and transcriptomic attributes between those who would survive and those who would succumb to COVID-19. We identified mRNA metabolism, RNA splicing, and interferon signaling pathways as key host responses overactivated by patients who would not survive. Those pathways are prime drug targets to reduce mortality of critically ill COVID-19 patients leading us to identify Tacrolimus, Zotatifin, and Nintedanib as three strong candidates for treatment of severely ill patients at the time of hospital admission.

**Teaser:** Epigenetics distinguishes COVID-19 survivors already at hospital admission: lessons for drug repurposing.

## INTRODUCTION

Coronavirus Disease 2019 (COVID-19) caused by severe acute respiratory syndrome-related coronavirus-2 (SARS-CoV-2) is characterized by heterogeneous clinical outcomes of infection ranging from absence of clinical symptoms to death. Intense research has resulted in the generation of highly effective vaccines for the prevention of COVID-19 related hospitalization and death (*1–5*). However, considering the ongoing circulation of the virus, the appearance of new variants with potential for escape of vaccine-induced immunity, the emergence of vaccine breakthroughs and wide-spread vaccine hesitancy, treatment of severely symptomatic COVID-19 patients remains a clinical challenge. Unfortunately, the success of these vaccines has not been paralleled by the development of efficacious drugs for the treatment of COVID-19 and there is an urgent need for effective therapies to treat severely ill COVID-19 patients. Treatment options include antiviral drugs (e.g. remdesivir), which have, so far, demonstrated only a discrete utility in the non-severe setting, or host-directed therapies. Of the latter, dexamethasone is currently recommended for patients who require supplemental oxygen, yet mortality rates remain in excess of 20% among critically ill patients (*6*). Therapeutic anticoagulation may reduce mortality in those with mild or moderate disease (*7*), while anti-IL6 antibodies (e.g. tocilizumab, sarilumab) can be added to steroid treatment for patients with severe manifestations and inflammation from SARS-CoV-2 (*8*). Remdesivir is currently approved by the Food and Drug Administration for treatment of COVID-19 patients (*9*), yet it has not been shown to improve mortality in patients hospitalized with COVID-19 and is not recommended by the WHO (*10*). A clinical trial in Spain showed that cyclosporine reduced mortality in hospitalized severe COVID-19 patients (*11*). Since the development of new antiviral drugs is a costly and time-consuming activity, drug repurposing has been the main avenue for discovery of treatment options of COVID-19 patients (*12, 13*).

Efforts of repurposing have focused on scanning key pathways of viral replication and host responses as possible drug targets. Protein-protein or protein-RNA interaction screens have not only provided exciting insight into the basic mechanisms of the SARS-CoV-2 life cycle but have also identified a multitude of candidate drugs for possible COVID-19 therapy (*14–18*). Similarly, artificial intelligence and computational modelling have been used to identify candidate repurposed drugs (*19–21*). Since results of these studies strongly rely on cellular assays, it is difficult to know how these findings will translate to patients. Patient-centric approaches have been focused on high resolution studies of immune processes and host responsiveness of hospitalized patients. These studies have defined key host responses that drive COVID-19 pathology and severity, and have pinpointed the type 1 interferon response as critical regulator of COVID-19 severity (*22–28*). In addition, these studies have identified single cell features, cell types and gene markers associated with the degree of COVID-19 pathology (*28–31*). However, these studies were primarily concerned with understanding COVID-19 pathophysiology and generally did not pursue a drug-repurposing strategy.

Given the spectrum of clinical COVID-19 presentations, it seems highly plausible that different drugs are needed for different disease stages. In our study, we have focused on critically ill patients for whom life-saving treatment remains an urgent unmet need. We reasoned that by contrasting critically ill patients who recover with those who perish, we would be able to identify host response pathways segregating between these two outcomes. Such pathways could then be pharmacologically targeted through enhancement-or suppression-based strategies. We enrolled patients with the same severe clinical WHO ordinal scale (OS=7) at hospital admission and followed these patients for 15 days post admission. Of the seven enrolled patients, three died of COVID-19 while four recovered and were discharged. We contrasted the transcriptome and epigenetic landscape of immune cells of the two patient groups at admission as well as at day 5 and 15 post admission as a follow-up. We found that disease evolution and survival are mainly characterized by changes in cellular composition with relatively modest impact on the transcriptome of key effector cells at later stages of the disease. In contrast, at the time of hospital admission, we detected significant transcriptional changes in key molecular pathways that are associated with epigenetic changes in monocytes. Of importance for the objective of the present study, at a time when the patients were of the same clinical severity, we identified the spliceosome and RNA metabolism as main targets for repurposed drugs to be deployed at the earliest point of hospitalization of critically ill patients.

## RESULTS

### Multi-omics profiling of severely ill hospitalized COVID-19 patients

We enrolled seven treatments naïve hospitalized patients with COVID-19 (P1 to P7) at the McGill University Health Centre, Montréal, Québec, Canada (Table 1). Among the seven patients, three were females and four were males. The average time from onset of symptoms to admission was 6 ±4.5 (SD) days and the average time from diagnosis to admission was 3 ±3.2 days. At admission, all patients were classified with an OS of 7 in the WHO Clinical Progression Scale (*32*). Three patients (1 female, 2 males) succumbed to the disease at days 8, 16 and 35 post hospital admission. For all patients, peripheral blood mononuclear cells (PBMC) were obtained by phlebotomy followed by standard density gradient centrifugation at three time points: at admission as well as five days (d5) and 15 days (d15) post recruitment (for surviving patients). This design allowed us to follow disease evolution from the same clinical reference point and to identify the epigenomics and cellular factors associated with COVID-19 disease evolution (Figure 1). Of importance, the retrospective classification of patients at admission in survivors and non-survivors permitted us to identify early changes in the overall immune status as high value targets for pharmacological intervention to block poor outcomes including death.

**Figure 1.**
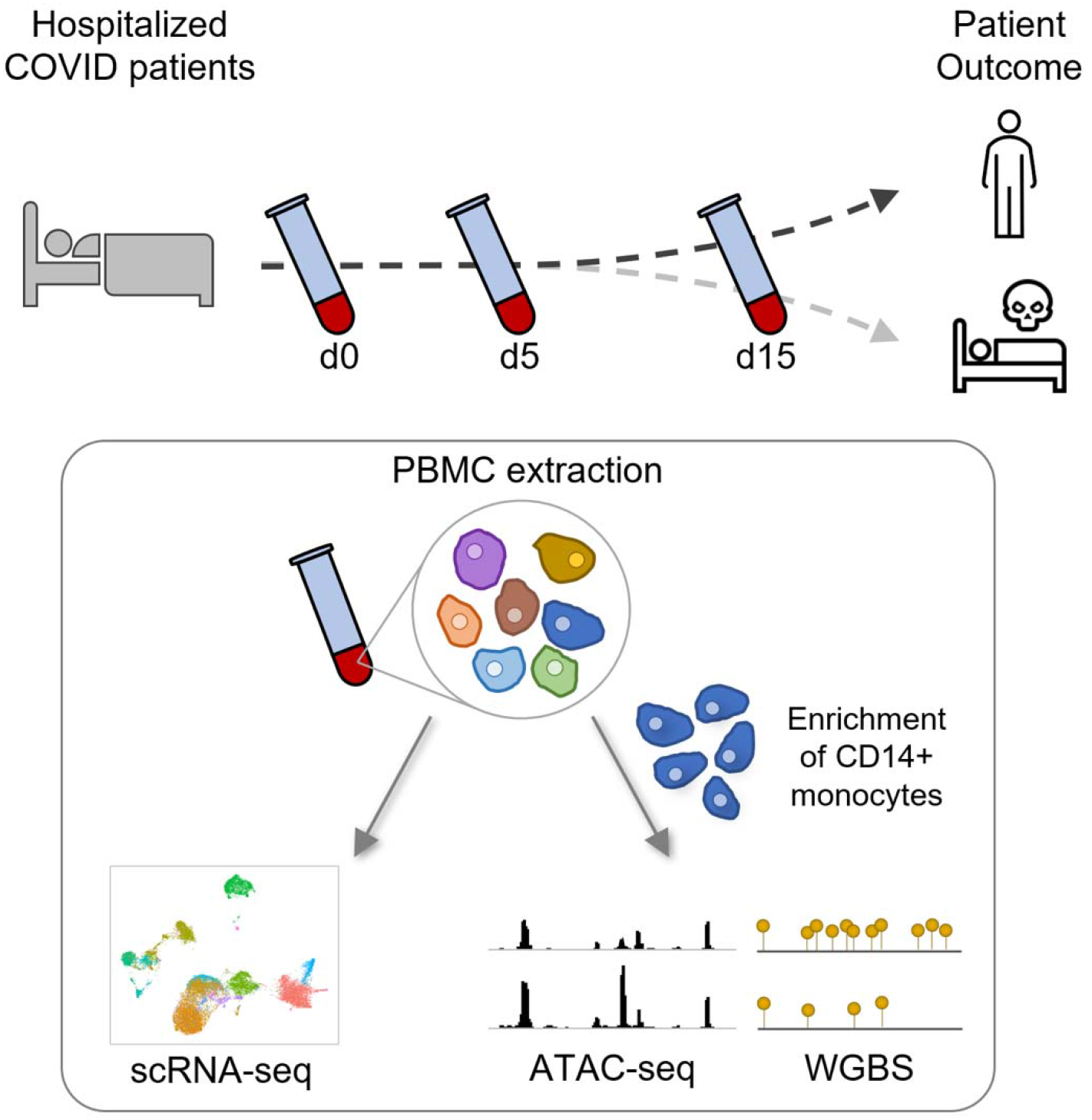
Study design summary. Blood samples were collected from severely ill COVID-19 patients at the time of hospitalization (d0) and at follow-ups (days 5 and 15) during their stay in the critical care unit. PBMC were isolated from the blood and captured for scRNA-seq. CD14+ monocytes were also enriched from PBMC for epigenetic analyses: chromatin accessibility by ATAC-seq and DNA methylation by WGBS.

**Table 1:**
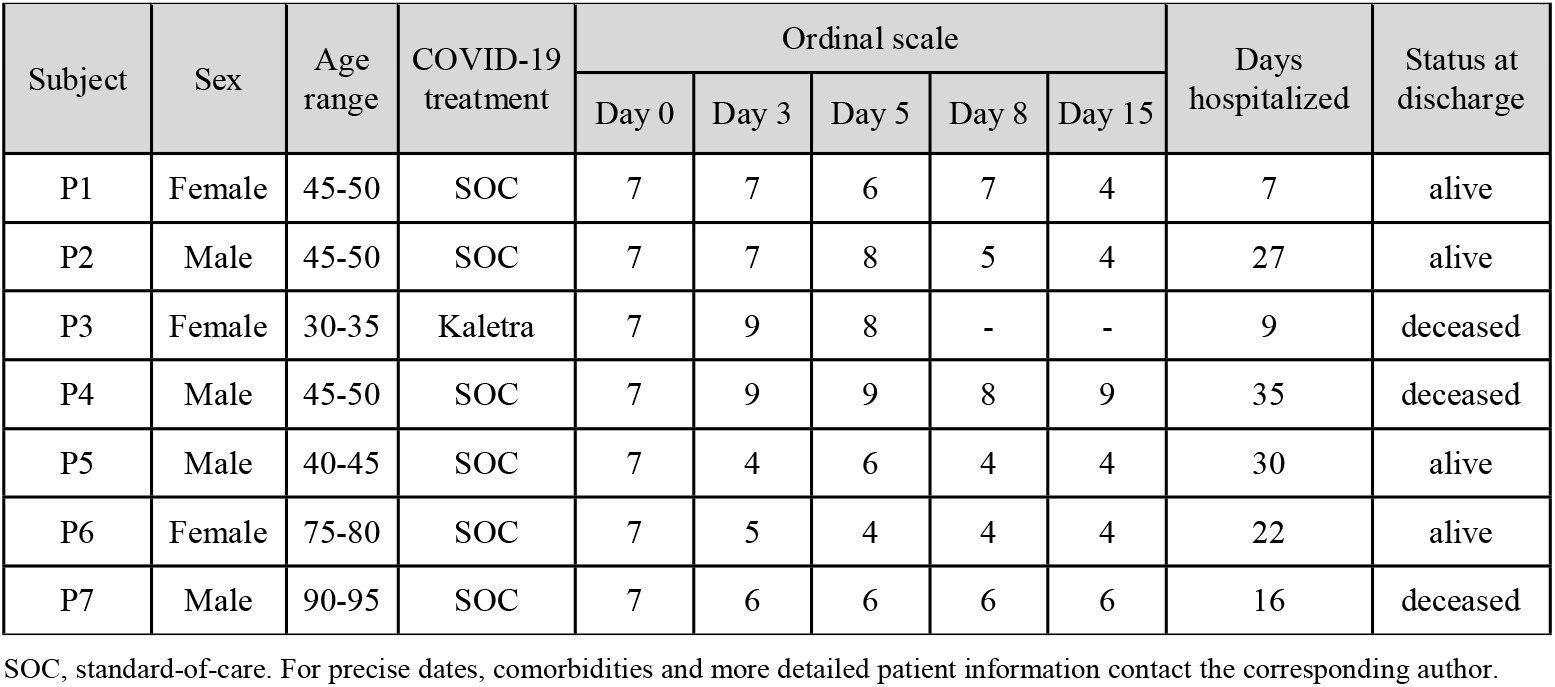
Patient demographics

| Subject | Sex | Age range | COVID-19 treatment | Ordinal scale |  |  |  |  | Days hospitalized | Status at discharge |
| --- | --- | --- | --- | --- | --- | --- | --- | --- | --- | --- |
|  |  |  |  | Day 0 | Day 3 | Day 5 | Day 8 | Day 15 |  |  |
| P1 | Female | 45-50 | SOC | 7 | 7 | 6 | 7 | 4 | 7 | alive |
| P2 | Male | 45-50 | SOC | 7 | 7 | 8 | 5 | 4 | 27 | alive |
| P3 | Female | 30-35 | Kaletra | 7 | 9 | 8 | - | - | 9 | deceased |
| P4 | Male | 45-50 | SOC | 7 | 9 | 9 | 8 | 9 | 35 | deceased |
| P5 | Male | 40-45 | SOC | 7 | 4 | 6 | 4 | 4 | 30 | alive |
| P6 | Female | 75-80 | SOC | 7 | 5 | 4 | 4 | 4 | 22 | alive |
| P7 | Male | 90-95 | SOC | 7 | 6 | 6 | 6 | 6 | 16 | deceased |

SOC, standard-of-care. For precise dates, comorbidities and more detailed patient information contact the corresponding author.

### Change in cellular components is associated with disease severity

We used a single-cell RNA sequencing (scRNA-seq) approach to understand the cellular composition and transcriptional status of PBMC following hospitalization and compared the results with those obtained for six healthy controls. Following quality filtering and doublet removal, a total of 105,851 cells were integrated and then clustered using a uniform manifold approximation and projection (UMAP) analysis. All samples were first regrouped into 3 major PBMC populations, namely B cells, Myeloid cells, and T+NK lymphocytes (Figure 2A). Cells from COVID-19 patients and cells from six healthy controls were homogeneously represented among the 3 clusters (Figure 2B). Clustering was confirmed by the expression of cell specific surface antigens (Figure 2C) and expression of known cell lineage marker genes (Figure 2D). We ran sub-clustering analysis for each of the 3 main lineages to confidently identify sub-populations. The T+NK lineage could be divided into 12 sub-clusters (Suppl. Figure 1), the B cells cluster into 4 sub-clusters (Suppl. Figure 2), and the Myeloid cell cluster into 7 sub-clusters (Suppl. Figure 3), which also included the plasmacytoid dendritic cells (pDC) and some hematopoietic stem and progenitor cells. Cell-type specific gene markers were extracted for every sub-cluster to identify the corresponding cell population.

**Figure 2.**
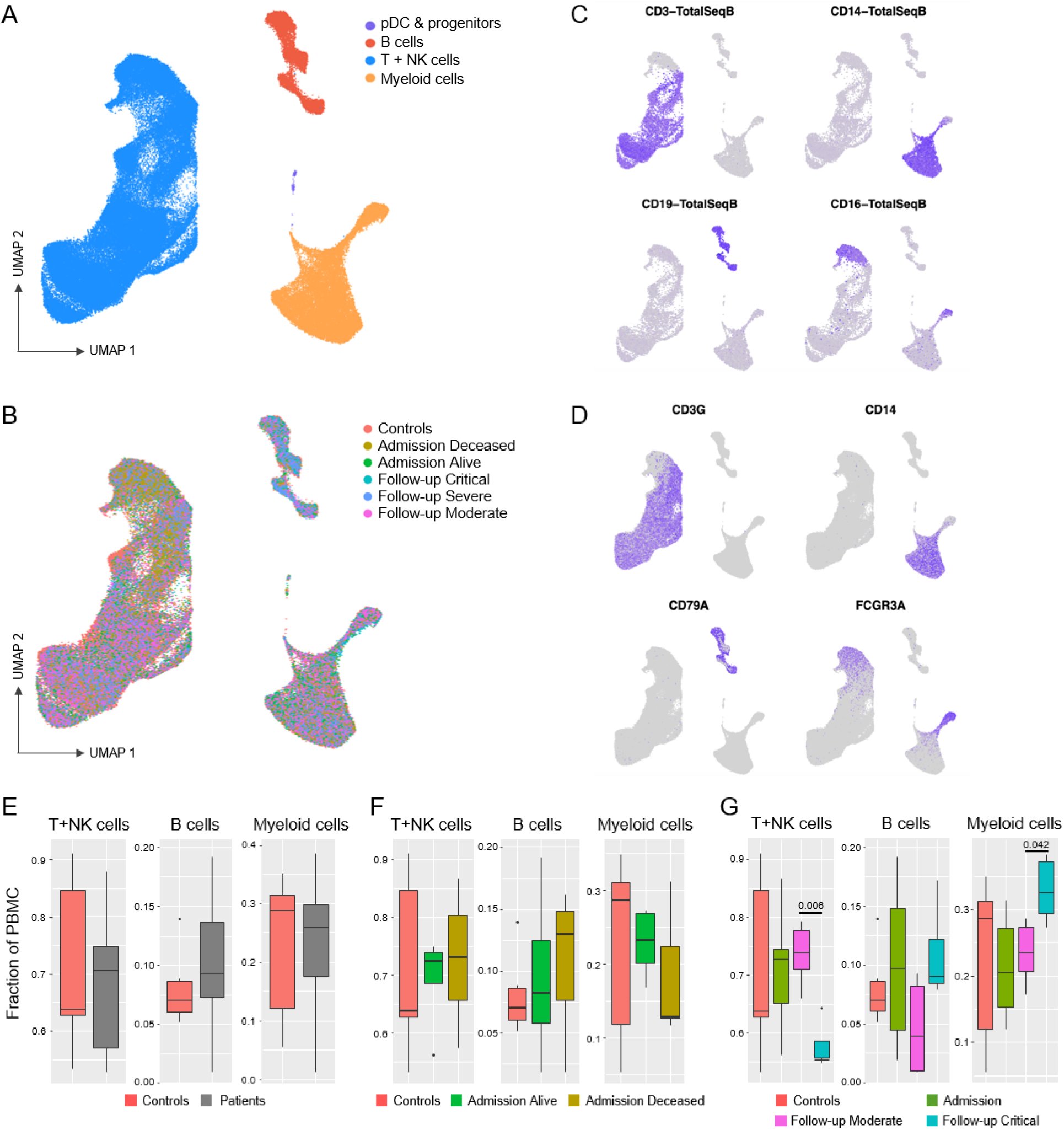
Cellular composition of PBMC from severely ill hospitalized COVID-19 patients and controls. **(A)** UMAP visualization of PBMC from 7 COVID-19 patients (18 samples obtained at days 0, 5 and 15 of admission) and 6 healthy controls identified four major PBMC lineages. **(B)** Cells are coloured according to the clinical classification of COVID-19 patients and healthy controls. **(C)** Visualisation of four cell surface lineage specific markers across cell clusters using TotalSeqB antibodies on control samples reveals T cells (CD3), CD14+ monocytes (CD14), B cells (CD19), and CD16+ and NK cells (CD16). **(D)** Cells are coloured according to the expression of the lineage marker genes *CD3G* (T cells), *CD14* (CD14+ monocytes), *CD79A* (B cells) and *FCGR3A* (CD16+ monocytes and NK cells). **(E-G)** Box and whisker plots for the proportion of cells in each major lineage compared to the overall number of cells in the sample for T+NK, B and Myeloid cells among clinical groups. The PBMC lineage proportions are shown for healthy control samples (red) versus **(E)** all COVID-19 samples (cyan); **(F)** samples collected from COVID-19 patients at admission and retrospectively classified as Deceased (blue) or Alive (green); and **(G)** samples at days 5 and 15 for patients classified as displaying Moderate (cyan) or Critical (purple) clinical symptoms. Q-values for pairwise t-test comparisons (Admission - Deceased vs Alive; Follow-up - Critical vs Moderate) are provided when significant.

PBMC lineage proportions were compared between patients and controls, as well as classes of patients (Figure 2E-G). In the global comparison, we did not detect significant differences in proportions of T-cells, B-cells and Myeloid cells between healthy controls and all severe COVID-19 samples (Figure 2E). Similarly, when patients were retrospectively grouped according to subsequent recovery (“Alive”) or death (“Deceased”), we failed to detect significant differences in PBMC lineage proportion for the corresponding day 0 samples (Figure 2F). In contrast, we observed a significant change in major lineage proportions when we pooled all samples according to their clinical status (i.e. Critical vs Moderate according to the OS score) at follow-up (day 5 + day 15; Figure 2G). This suggested a strong impact of developing COVID-19 severity on PBMC proportions. Critically ill patients showed a significant reduction of T cells (*p*-value = 0.006) and a significant increase of Myeloid cells (*p*-value = 0.04). These are accounted by the changes in specific cellular subsets, where (i) the decrease in T cell numbers was driven by a specific reduction of naive (*p*-value=0.004) and central memory (Tcm; *p*-value=0.04) CD4+ T cell populations (Suppl Figure 4A); (ii) B cell sub-populations (Suppl Figure 4B) remained unchanged and confirmed the absence of association with disease severity; and (iii) the increase in Myeloid cells was driven by a specific expansion of CD16+ monocytes (*p*-value=0.03; Suppl Fig 4C).

### Transcriptional expression profile as a marker of the COVID-19 prognosis

To gain further insight in the molecular pathogenesis of COVID-19 severity, differential gene expression (DGE) analyses were conducted for PBMC populations. We excluded the pDC, cDC2 and progenitor populations from this analysis since the total number of identified cells per population was too low (<300) to generate confident results. For comparisons, we employed the same grouping as used for the analysis of lineage proportions, i.e COVID-19 patients “Deceased vs Alive” at day 0, “Critical vs Moderate” at follow-up (see Suppl. Table1 for detailed DGE results per lineage). All significantly differentially expressed genes (adjusted *p*-value ≤ 0.05) were used for GO enrichment analysis, with the over-expressed and lower-expressed genes analyzed separately. The significantly enriched GO terms (adjusted *p*-value ≤1e-5 and enriched for at least 5 differentially expressed genes) in at least one cell population were selected and re-examined in every population for the above contrasts. The corresponding results are summarized in the heatmap in Figure 3A and detailed in Suppl. Table 2. Strikingly, despite no differences in WHO clinical score at admission, we observed more transcriptional changes and thus more enriched pathways for the “Decreased vs Alive” comparison at day 0, than for the “Critical vs Moderate” contrast at later time-points (Figure 3A). Based on GO enrichment at admission, we identified three groups of cell populations: high responder cells (HR-cells: CD14+ and intermediate monocytes), moderate responder cells (MR-cells : CD16+ Monocytes, mature B cells, cytotoxic NK CD56dim, and various T cell subsets: NKT-like, Cytotoxic, exhausted (Tex), central memory (Tcm), effector memory (Tem), and Naive CD4+ T cells), and low responder cells (LR-cells: NK CD56bright considered to be high cytokine producers, Activated T cells, δT, regulatory T cells (Treg), Naive T CD8+ and Naive B cells) with no significant pathway enrichment in the latter cells (Figure 3A, top of heatmap).

**Figure 3.**
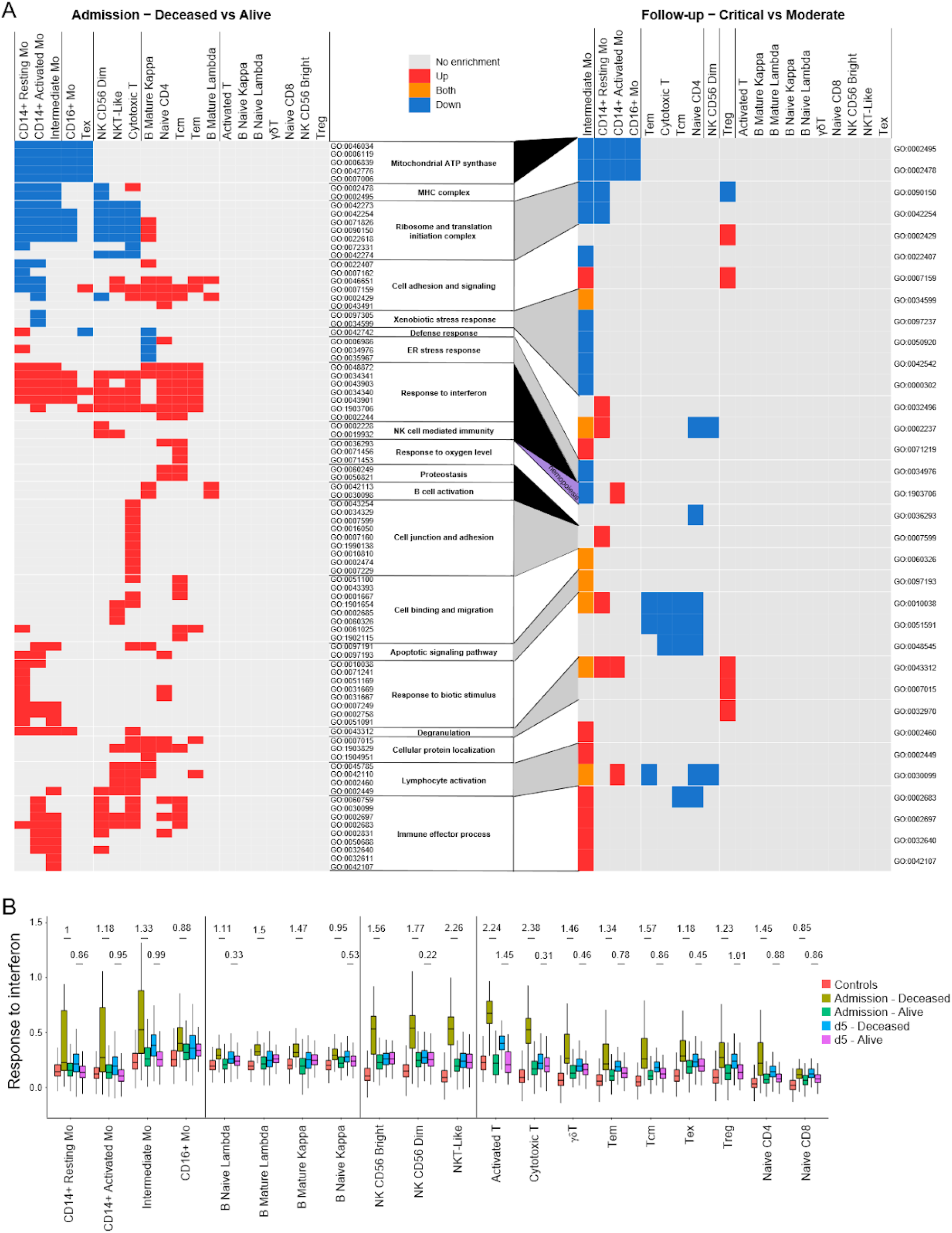
Transcriptional variations in cell populations of severely ill COVID-19 patients. **(A)** GO-term enrichment analysis for differentially expressed genes at admission between the “Deceased vs Alive” patient groups (left heatmap) and between the “Critical vs Moderate” COVID-19 patients at follow-up (right heatmap). Heatmap boxes are colored based on the enrichment observed for the hierarchical level 5 of the GO Biological-Process sub-ontology tree in the corresponding cell lineage: Grey for no significant enrichment; Blue, Red and Orange, respectively, for significant (q-value <= 0.05) enrichment of down-regulated, up-regulated and both up- and down-regulated genes. Cell population enrichment clusters have been generated using a complete-linkage clustering approach independently for both heatmaps resulting in a different order of lineages. GO categories only enriched at admission are highlighted in black between the two heatmaps, whereas the GO category highlighted in yellow (Hemopoiesis) represents the only category enriched only at follow-up. GO categories enriched at both admission and follow-up are successively highlighted in white and grey for easier visualization. **(B)** Evolution of the transcriptional expression for the *Response to Interferon* GO-term. Differentially expressed genes enriched in the GO-term in (A) were used to generate a Module Score representing the overall expression level for term genes in the cell lineage indicated at the bottom of the graph. Module scores shown were derived at admission and the day5 time-points for the “Deceased” and “Alive” COVID-19 patients separately. Control samples from healthy subjects are included as reference. Significance of differences between “Deceased” and “Alive” COVID-19 patients was tested by comparing the corresponding Module Score distribution in every cell subset using a Wilcoxon rank-test approach (q-value <= 0.05). To provide a sample-size free evaluation of the difference in expression, Cohen’s d effect size estimation is shown for every significant variation. Transcriptional evolution graphs for the other enriched GO categories are provided in Suppl. Figure S5. Tcm: Central memory T cells; Tem: Effector memory T cells; Tex: Exhausted T cells; Treg: Regulatory T cells; gdT: gamma delta T cells; B cells: either naive or memory cells expressing either the kappa or lambda light chain.

To further investigate expression changes of genes enriched in GO terms, we considered enriched genes as Modules and calculated Module Scores for each cell population to facilitate comparisons between subject groups. Module Scores were also computed for the “Deceased vs Alive” groups at day 5 in order to observe how these transcriptional changes progress along the disease evolution. At admission, the highest Module Score was obtained for genes in “Response to Interferon” identifying this as the most upregulated term, but only in patients that went on to die from COVID-19 (Figure 3B). In both HR- and MR-cells, this was accompanied by the increased expression of genes involved in “Immune Effector” processes suggesting that exacerbated immune responses and type I interferon signalling in HR- and MR-cells were linked with poor prognosis for patients (Suppl. Figure 5I).

Overall, the Module Scores highlighted that Activated/Cytotoxic T cells and NK cells at admission are globally more activated than other lymphocyte populations in patients with poor prognosis, with strong induction of genes in the “Immune effector process”, “NK cell-mediated immunity”, and “Lymphocyte activation” terms (Suppl. Figure 5). Similarly, T and NK cells exhibited increased expression of genes enriched in “Cell junction and adhesion” and “Cell binding and migration” terms. In addition, CD4+ naive T cells and Tcm cells displayed strong expression of genes enriched in “Apoptosis”, “Response to oxygen levels”, “Proteostasis” and “ER stress response” terms. Interestingly, both naive CD4+ and Tcm subpopulations underwent a strong reduction in numbers by day 5 and 15 post admission (Suppl. Figure 4). In monocytes, poor prognosis at admission was also linked with a reduced expression of genes involved in the “Mitochondrial ATP Synthase” (mainly genes of the complex V), the “MHC Complex” and “Ribosome and Translation Initiation” terms. Finally, in mature B cells we observed increased expression of genes enriched in the “B cell activation” term in non-survivors, consistent with a targeted response to SARS-CoV-2 in specific lymphocytes sub-populations.

We next compared the gene expression between patients who exhibited “Critical” vs “Moderate” symptoms at follow-up. Overall, transcriptional differences were small and correlated with the “deceased” prognosis at admission. A majority of terms that had shown significant enrichment at admission between “Deceased vs Alive” patients were not detected in this comparison (Figure 3A). However, we still detected significant differences between gene expression Module Scores on day 5 especially in “Response to Interferon”, “Immune Effector Process”, and “Lymphocyte Activation” terms (Figure 3B; Suppl. Figure 5). Importantly, we noticed that monocytes expressed most of the remaining transcriptional differences between admission and the later stages of the disease. Additionally, the Module Score analysis of “Deceased vs Alive” patients at follow-up showed lower or null effect size when compared to those observed at admission consistent with the suggestion of a reduced transcriptional impact at later stages of the COVID-19 disease.

The reduced transcriptional differences contrasted with the substantial differences of PBMC lineage proportions that were only observed at later stages of hospitalization. This suggested that variations in transcriptional activity, and the accompanying epigenomics changes, mostly occurred at an early stage of COVID-19 disease, dictating how the disease will evolve in terms of severity and final outcome.

### The epigenetic profile of CD14+ monocytes correlates with poor prognosis for COVID-19

Given that monocytes had the largest number of transcriptional differences in our scRNA-seq, we sought to analyze their chromatin accessibility using the Assay for Transposase-Accessible Chromatin (ATAC-seq) on CD14+ monocytes enriched from PBMC of hospitalized COVID19 patients over the two weeks follow-up (Figure 1). We observed an increase in the fraction of reads in peaks (FRIP), a surrogate of cell quality and genomic stability, between day 0 and days 5 and 15 (Suppl. Figure 6A). Genomic stability negatively correlated with an increase of the clinical OS, with critically ill cases presenting lower FRIP (Suppl. Figure 6B). The high background resulting from the chromatin instability in patient monocytes restricted the comparison of genomic regions between phenotypic groups. Therefore, we were only able to test for differential accessible chromatin (DAC) in 13398 genomic regions where reads in peaks surpassed the background noise.

We contrasted the epigenetic profile of CD14+ monocytes from the “Deceased” patient group with the one from the “Alive” group at day 0 and at follow-up (days 5 + 15). Of the 13398 chromatin regions tested, 959 were DAC at admission and 407 were DAC at follow-up (Figure 4A; Suppl. Table 3). Interestingly, 222 of the 407 DAC regions (54.5%) at follow-up overlapped with DAC regions at day 0 (Figure 4B and Suppl. Table 3). Among the “Deceased” group, the proportion of regions with sustained chromatin changes at follow-up was higher for less accessible regions (62.2%) compared to more accessible regions (32%; Fisher Exact Test, *p*=1.1e-7; Figure 4B). Hence, while chromatin accessibility between the two groups became less pronounced, there was still a trend to maintain accessibility status of DAC regions between admission and follow-up (Figure 4C).

**Figure 4.**
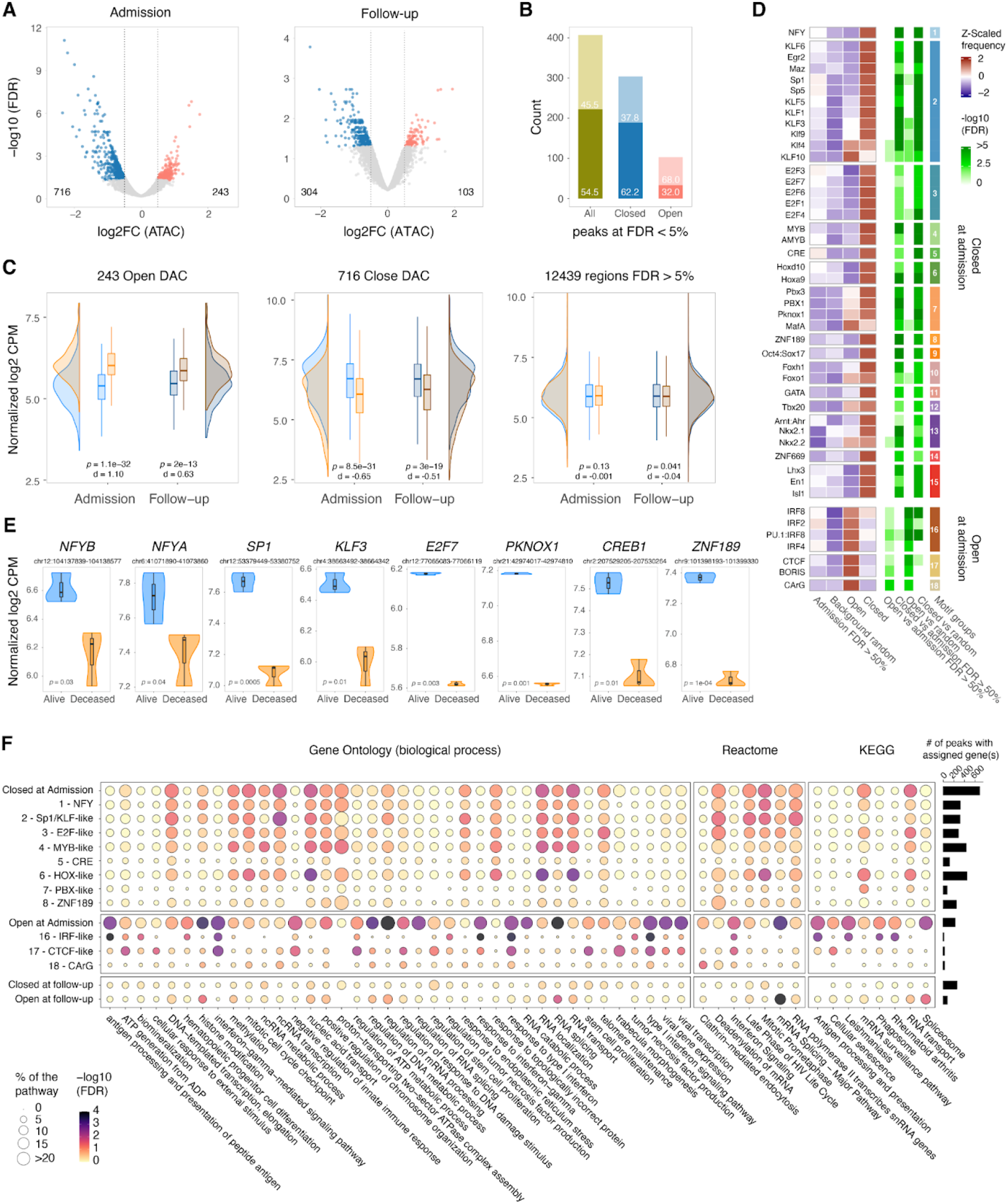
Monocyte chromatin accessibility among COVID-19 patients. **(A)** A total of 13,398 genomic regions identified as accessible by ATAC-seq were tested for differential accessible chromatin (DAC) and plotted according to their log2 fold change and corresponding adjusted *p* value for the “Deceased vs Alive” comparison at day 0 and follow-up (days 5 and 15). More closed and more open chromatin regions in “Deceased” patients at a false discovery rate (FDR) < 5% and absolute log2 fold change (log2FC) > 0.5 are shown in blue and red, respectively. **(B)** Intersection of DAC regions at admission and follow-up for the “Deceased vs Alive” group comparisons. The proportions of the 407 significant DAC regions at follow-up that were also significant at d0 are shown in darker shade. The overlap for open DAC is indicated by the red bar while the overlap for closed DAC is indicated by the blue bar. **(C)** Plots indicating the distribution of chromatin states for regions significantly more open (left), more closed (center) and not significantly different at FDR >5% (right) for the “Deceased vs Alive” comparisons. The normalized log2 count per million (CPM) corresponding to accessibility levels for each of the chromatin regions denoted at the top was plotted in the y-axis and summarized as density and box plots for the admission and follow-up samples. Blue shading indicates the chromatin state for the “Alive” group and orange shading indicates the “Deceased” group. A Cohen’s d value was used to estimate the effect size and a wilcoxon test to assess the significance of the differences of the distributions between the “Deceased” and “Alive” patient samples. **(D)** Motif enrichment analysis for 959 DAC regions between “Deceased vs Alive” at Admission. Of the 682 motifs tested, 46 were significantly enriched in DAC regions of deceased patients at FDR <1% and are plotted as a heatmap. Of them, 39 motif groups were more frequently observed in DACs less accessible (Closed) in deceased patients, while seven motifs were observed more frequently in open DACs. A total of nine columns are identified at the bottom of the heat maps. The four columns on the left represent heatmaps of the Z-scaled motif frequency. The first two columns indicate motif frequencies in control groups including non-significant regions at FDR >50% at admission and 50 thousand random genomic regions with the same length distribution as significant DACs, followed by open and closed DACs at admission. Shades of red indicate higher frequency and shades of blue lower frequency for each motif. The four columns at center of the heatmap indicate the negative log10 FDR of the enrichment for the motif shown in the first four columns. In the last column, transcription factors are grouped according to the homologies of their respective binding sites (details in Suppl. Figure 7). **(E)** Suppressed promoter accessibility in the “Deceased” patient group for eight TFs with motifs enriched in closed DAC regions at admission. Chromosomal locations of the peaks are shown below the TF name. Normalized log2 read counts (as counts per million, CPM) for promoter peaks are plotted on the y-axis for the “Deceased” and “Alive’’ groups. The corresponding chromatin tracks are shown in Suppl. Figure 8. **(F)** Gene ontology and pathway enrichment analyses for DAC regions at admission and follow-up (day 5 and 15) for the “Deceased vs Alive” contrast. Genes assigned to DAC regions (details in methods) or DACs stratified by motifs with most significant FDR p-values on Figure 4D were used to interrogate the KEGG and Reactome pathways, and the GO biological process terms databases. Non-redundant pathways and GO terms presenting an FDR < 5% and at least five genes in one of the tested conditions are plotted, with the bubble size representing the percentage of genes in a given pathway or term, and the colour shades indicating the negative log10 FDR. Representative examples of TF with enriched motifs in closed or open DAC regions are listed to the left of the graph. The number of DACs corresponding to each peak category on the left is shown as a bar plot on the right.

To understand the biological mechanisms tagged by epigenetic differences in the “Deceased vs Alive” comparison, we performed DNA binding motif enrichment for DAC regions at admission. We found motifs for seven transcription factors (TFs) enriched in chromatin regions of increased accessibility in “Deceased” patients (Figure 4D). Among these motifs was a set of interferon responsive factors (IRFs) known to regulate interferon responses, and CTCF which is involved in regulating transcription and chromatin architecture (Figure 4D and Suppl. Figure 7). An enrichment of motifs for 39 TFs was observed in regions less accessible in “Deceased” patients. As many of these TFs bind to homologous motifs, we summarized the TFs in groups by similarities in their binding sites (Figure 4D and Suppl. Figure 7). The groups most represented in less accessible regions included transcription factors with Sp1-like and E2F-like binding motifs. These TF families have multiple regulatory functions including cell cycle, differentiation, and chromatin remodeling, but are also associated with immune responses. Potentially as part of feedback loops, TF members of six out of the top eight motif groups had their promoter region also less accessible in “Deceased” patients (Figure 4E, Suppl. Figure 8).

Next, we performed gene ontology and pathway analyses using genes assigned to more open or closed DAC regions at admission and follow-up. Additionally, we stratified the analysis using genes assigned to DAC regions overlapping motifs for the leading TF of each group (Figure 4F). For DAC regions more open at admission, we observed the type-1 interferon response pathway enriched for both IRF-like and CTCF motifs (Figure 4F, Suppl. Table 4). Peaks with CTCF and IRF binding motifs were independent and assigned to different sets of genes that converged in type-1 interferon pathways (Suppl Table 4). Other terms and pathways tagged by more accessible chromatin included mRNA splicing and ATP metabolisms. Interestingly, the ATP metabolism pathways were mostly tagged by peaks with CTCF motifs, while spliceosome and RNA metabolic pathways were not specific to a TF motif group (Figure 4F). Conversely, in accordance with scRNAseq results, RNA transport and proton-transporter for ATP GO terms were more frequently observed in closed chromatin of deceased patients suggesting an unstable RNA metabolism and transport in CD14+ monocytes.

### Overlap of DAC with severe covid-19 with respiratory failure GWAS

In a previous genome-wide association study (GWAS) (*33*), two significant and three suggestive loci were associated with severe COVID-19 with respiratory failure. Out of these five GWAS loci, two overlapped multiple DAC regions for the “Deceased vs Alive” at admission contrast (Figure 5). The overlap between DAC regions and the GWAS loci was significantly not random (*p* = 0.01; Suppl. Figure 9). The most significant locus associated with hospitalization in the COVID-19 GWAS encompassed the *FYCO1* and *LZTFL1* genes on chromosome 3. We observed that the promoter region of these two genes is more accessible in the “Deceased” patient group, with the *FYCO1* promoter also maintaining increased accessibility in the follow-up (Figure 5). Interestingly, for the suggestive COVID-19 protective signal observed on chromosome 5, the promoter regions of two flanking genes *CEP120* and *CSNK1G3* were less accessible in the “Deceased” group at hospital admission and follow-up (Figure 5).

**Figure 5.**
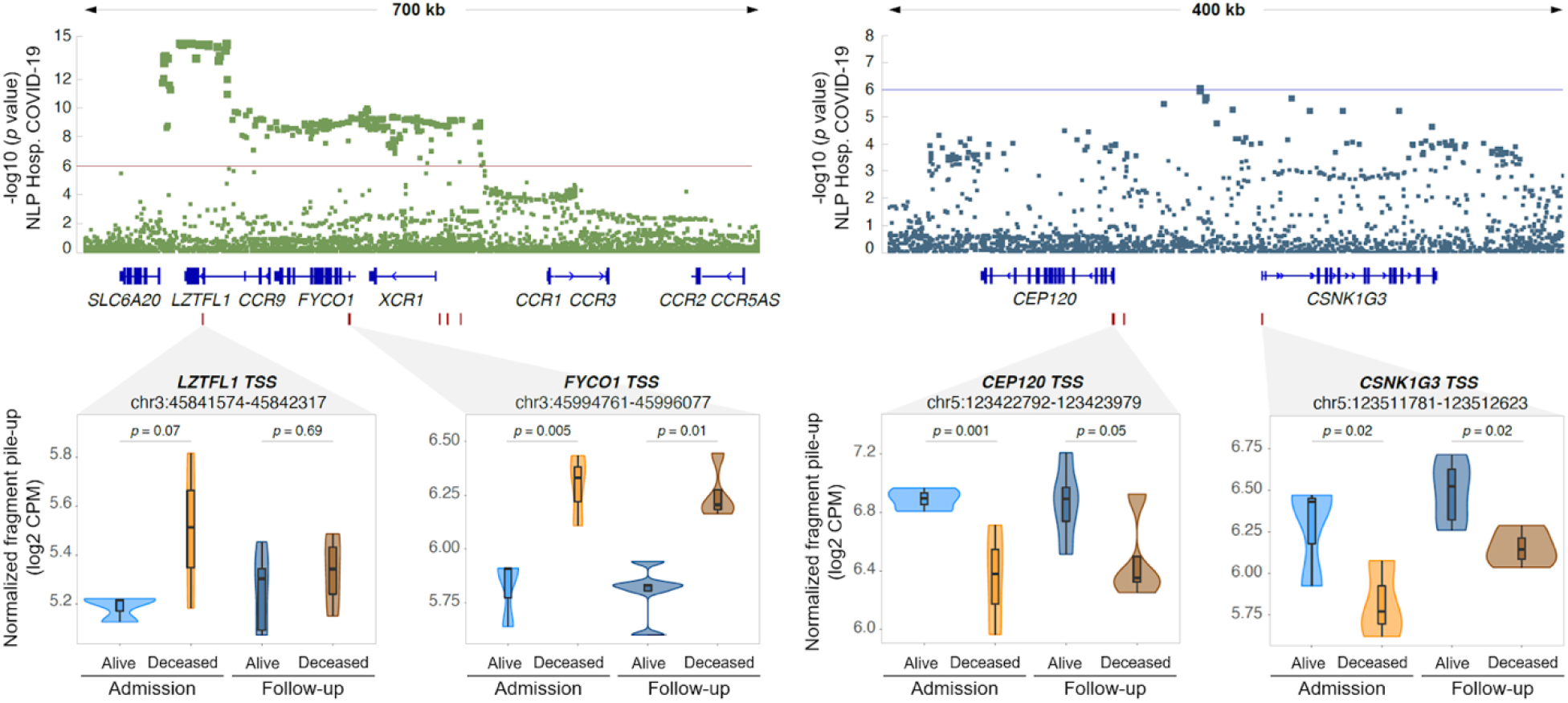
Differential open chromatin at genes located within GWAS loci for COVID-19 hospitalization. Two out of the five GWAS loci for severe COVID-19 with respiratory failure contain multiple DACs in monocytes from hospitalized COVID-19 patients (*33*). The y-axis indicates the evidence for association of SNPs (dots in the top panels) with severe COVID-19 as negative log10 p-value. The x-axis indicates the chromosomal location of SNPs with colocalizing genes shown below. A zoom-in at the four regions with DAC for the “Deceased vs Alive” contrast is shown in the lower part of the figure. Box plots with normalized quantification of pileup fragments in peaks at promoter regions for “Deceased” and “Alive” are shown as log2 count per million (CPM) for patient groups at admission and follow-up.

### Differences of DNA methylation between patient groups at admission

To further investigate changes in chromatin in CD14+ monocytes at admission, we performed whole genome bisulfite sequencing and compared 26,840,057 CpG loci between the “Deceased vs Alive” patient groups. From the 222,751 differentially methylated loci (DMLs) with a *p*-value < 0.05 (Suppl. Figure 10A), we discovered 6,259 differentially methylated regions (DMRs; Suppl. Figure 10B). Approximately half (48.5%) of the DMRs were hypermethylated in “Deceased” patients. While the majority of DMRs were located in genic and intergenic regions, 7.13% of DMRs were located in gene promoter regions (Figure 6A). Overall, DMRs were distributed across the genome (Figure 6B).

**Figure 6.**
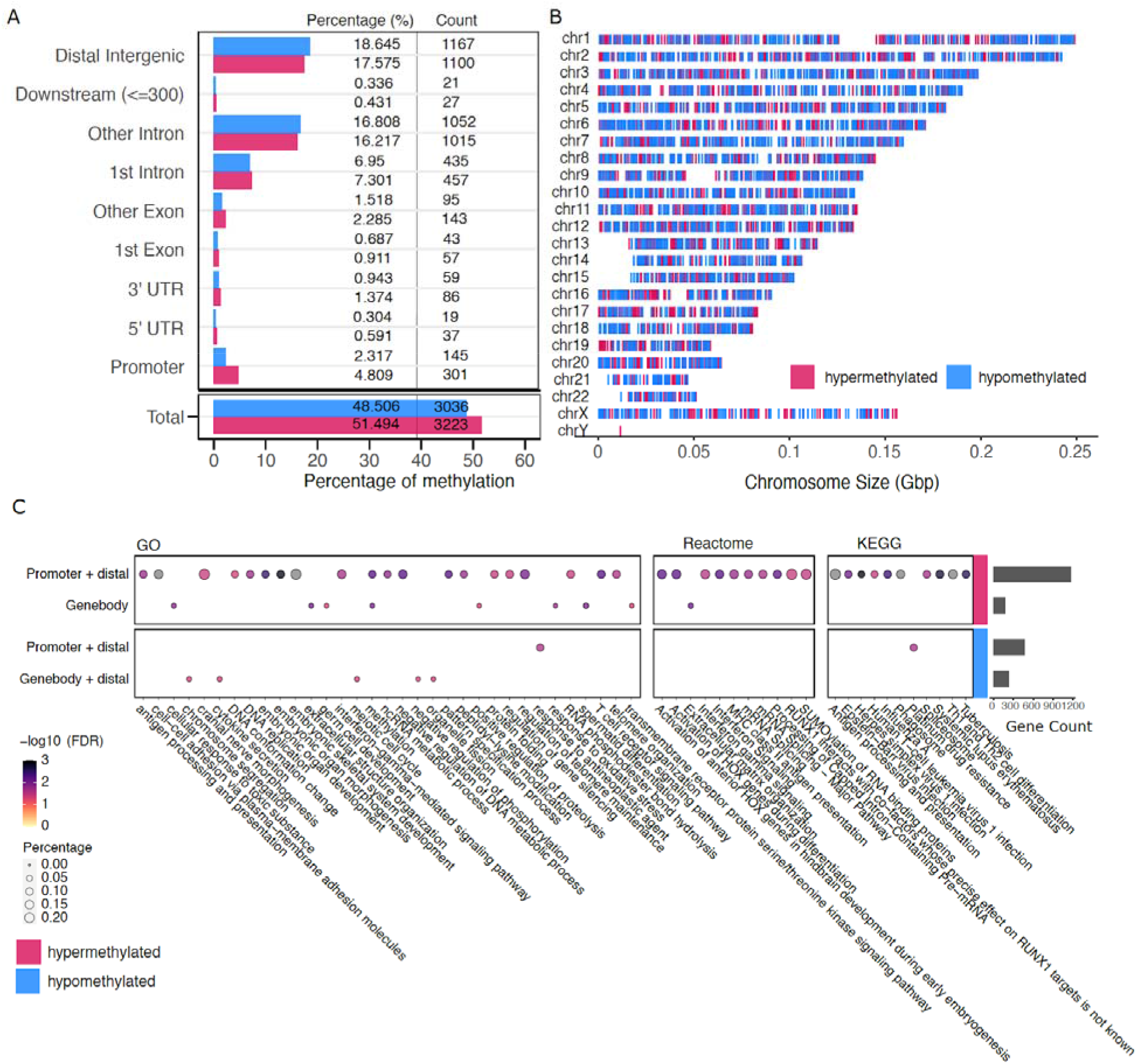
DNA methylation analysis of monocytes at admission. **(A)** Count and percentage of hyper (red) and hypo-methylated (blue) regions of Deceased vs Alive patients across annotated genomic regions. **(B)** Distribution of hyper-(red) and hypo-methylated (blue) regions across chromosomes. Each vertical line indicates a DMR. **(C)** Gene ontology and pathway enrichment analyses for DMRs at admission comparing “Deceased vs Alive” patients. Genes assigned to DMRs in promoters, gene bodies, and distal regions (details in method) were used to search the KEGG and Reactome pathways, and the GO biological process terms databases. Genes associated with hypermethylated DMRs are shown in the top two rows and hypomethylated DMRs in the bottom row. Non-redundant pathways and GO terms showing FDR <5% and with at least five unique genes and DMRs in one of the test conditions are plotted; bubble size represents the FDR and colour shading the percentage of genes with a corresponding significant DMR at FDR <5%. The total number of genes per group associated with DMRs used to interrogate the databases is shown in the bar plot on the right.

To evaluate the effect of these DMRs in COVID-19 pathogenesis, we performed a TF binding motif enrichment analysis as implemented for DAC regions and found no significant enrichment at FDR < 0.01. Next, we performed GO-term and pathway analyses based on genes tagged by promoter, distal and gene body DMRs. Using hypermethylated DMRs, located at promoters or tagging distal genes, we observed spliceosome-related pathways and antigen processing and presentation terms significantly enriched in deceased patients (Figure 6C, Suppl. Table 5). In addition, for DMRs in gene body, several pathways including the extracellular matrix organization were significant. Moreover, GO terms and pathways including cytokine secretion and negative regulation of phosphorylation were enriched in gene body and/or distal DMRs. No enrichment was found using hypomethylated DMRs located in the gene body and hypermethylated DMRs located in the gene body combined with distal DMRs.

A recent study showed that changes in gene expression in response to infections occur prior to detectable alterations in the methylome (*34*). Therefore, we hypothesized that methylation changes might increase only at advanced disease stages after the admission. Using the same “Deceased vs Alive” design applied to follow-up samples, we detected 491,208 DMLs (*p*-value < 0.05) (Suppl. Figure 10C). DMLs were then used to discover a total of 21,516 DMRs (Suppl. Figure 10D). The intersection between the DMRs found at admission and follow-up suggested that approximately 40% of DMRs were conserved during disease progression (Suppl. Figure 11A). When we conducted gene ontology and pathway analyses for the genes tagged by DMRs at follow-up, we observed 16 (25.6% from admission) pathways and GO-terms that were already enriched at admission and thus maintained during disease progression, including spliceosome-related pathways and positive regulation of telomere maintenance (Suppl. Figure 11B, Suppl. Table 6). Additionally, several pathways and GO-terms including chromatin modifications, RNA localization and modifications were observed exclusively at follow-up. Interestingly, these pathways were already observed altered using DAC regions at admission, suggesting delayed changes at the methylome.

### Data integration and identification of candidate drugs

We observed a high level of consistency for GO/Pathway terms between chromatin accessibility and gene expression. Specifically, in the “Deceased” patient group the majority of terms tagged by upregulated genes were likewise detected by increased chromatin accessibility (Figure 7A). The concordance of epigenetic and transcriptomic changes was often observed for individual genes. For example, genes such as *IFITM2, IFITM3*, *IFI6* and *ISG15* included in the “type-1 interferon signaling” term were upregulated in “Deceased” patients at admission and also displayed increased promoter accessibility (Suppl. Figure 12). As the disease advanced, both transcriptional and chromatin structure differences in the “Deceased vs Alive” contrast attenuated (Suppl. Tables 1 and 3). Significant differences in splicing-related pathways between “Deceased vs Alive’’ patients were observed at the gene expression, chromatin accessibility and DNA methylation levels (Figure 7A). Moreover, RNA Splicing related terms were among the most significant pathways detected by open DACs and hypermethylated DMRs at follow-up, suggesting that “Deceased” patients could not suppress splicing-related mechanisms (Figure 4F, 6C). The observation that reduced chromatin accessibility of mRNA splicing-related genes improved survival was also highlighted by corresponding transcriptomic changes (Figure 7A). Hospitalized patients who survived (“Alive” group) showed in monocytes, B, NK and T cells, a significantly lower expression for genes that are part of the mRNA Splicing pathway than patients who died (Figure 7B). Clearly, the identification by different assays of the same differentially activated pathways in cells from COVID-19 survivors and decedents implicates these pathways as determinants of prognosis, and thus supports the validity of these pathways as targets for pharmacological intervention.

**Figure 7.**
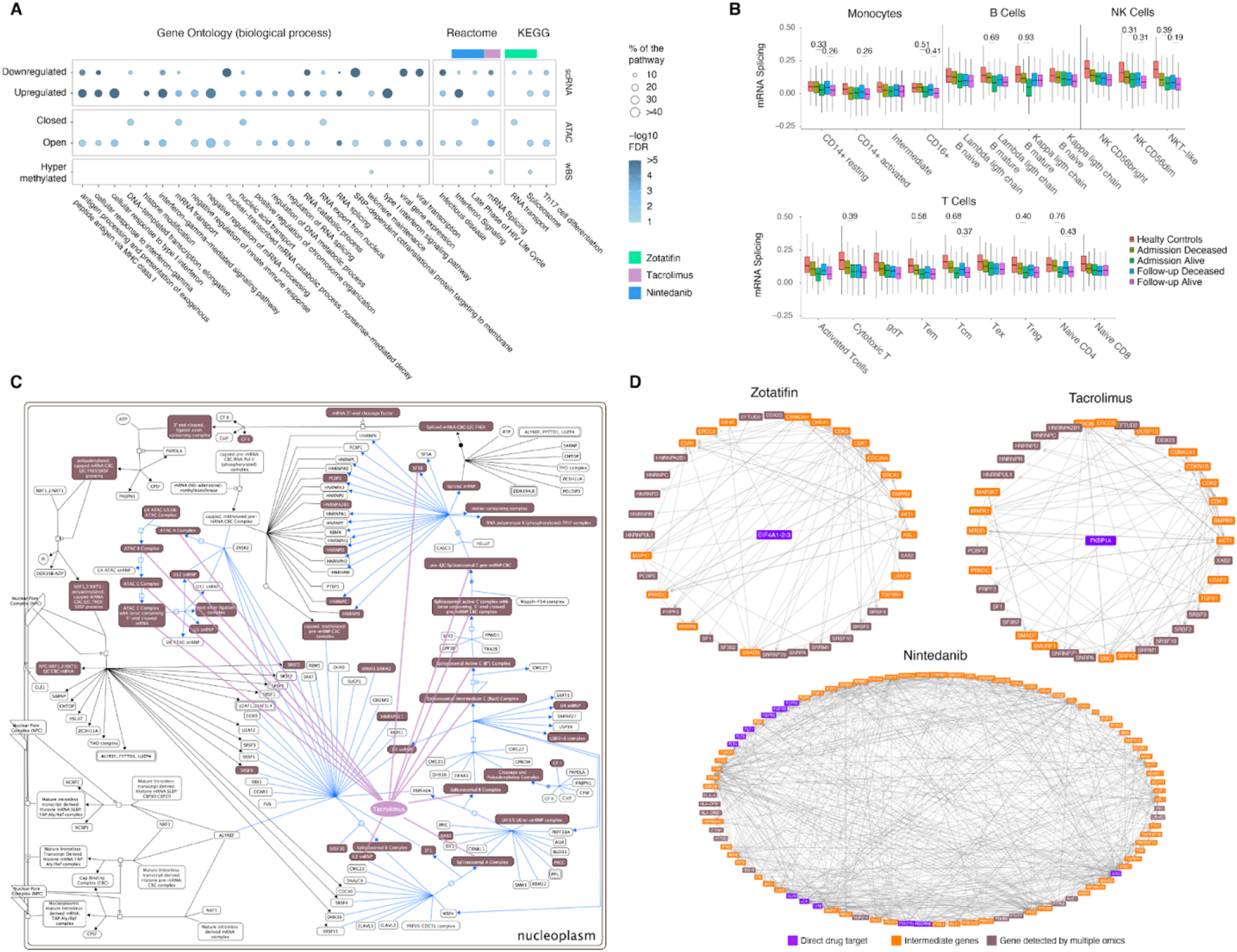
Candidate drugs to treat hospitalized COVID-19 patients. **(A)** Pathways and GO biological process terms identified by differential transcriptomic, chromatin accessibility, and/or DNA methylation are shown in the bubble plot. The bubbles indicate pathways or terms with more than 10 genes with a FDR < 5% (scRNA-seq and ATAC-seq) or FDR < 10% (WGBS). Colour shades of the bubbles represent the −log10(FDR) and the bubble size indicates the proportion of genes in the pathway or term that were significantly different between “Deceased” and “Alive” patient groups. The shared pathways and terms targeted by the Tacrolimus, Zotatifin, and Nintedanib drugs are shown at the top of the graph with colour shades corresponding to the drugs listed on the right of the graph. **(B)** Evolution of the transcriptional expression for the *mRNA Splicing.* Differentially expressed *genes enriched in the* Reactome’s *mRNA Splicing* pathway were used to generate a Module Score representing the overall transcriptional expression level for the cell lineage indicated at the bottom of the graph. Module scores shown were derived at the D0 and D5 time-points. For each time point, the Module Score was estimated for the “Deceased” and “Alive” severely ill COVID-19 patients separately. Control samples from healthy subjects were included as reference for transcriptional expression levels. Significance of differences in the transcriptional expression levels between “Deceased” and “Alive” severely ill COVID-19 patients was tested by comparing the corresponding Module Score distribution in every group of cell subsets using a Wilcoxon rank-test approach (q-value <= 0.05). To provide a sample-size free evaluation of the difference in expression, Cohen’s d effect size estimation is shown for every significant variation observed. **(C)** Tacrolimus interacts with mRNA splicing hubs identified by significant transcriptomic and epigenetic changes between patients at admission who will succumb to COVID-19 and those who will recover. The mRNA splicing portion of the Processing of Capped Intron-Containing Pre-mRNA pathway is indicated by light blue lines. Genes and hubs tagged by both transcriptomic and epigenetic changes are highlighted by a brown shade. **(D)** Protein-protein interaction network targeted by candidate drugs. The proteins targeted by the candidate drugs were accessed in DrugBank and intersected with OmnipathR protein-protein interaction databases. Proteins targeted by the drugs (Zotatifin, Tacrolimus and Nintedanib) are shown in purple. The shortest interaction between protein drug targets to proteins encoded by genes identified by our study marked in brown via intermediate genes marked in orange are linked by light grey lines. For Zotatifin and Tacrolimus, proteins in the mRNA splicing pathway, and for Nintedanib proteins in the Interferon Signaling pathway identified in our study as significant DGE or DAC are shown in brown.

We searched for existing drugs interacting with pathways and GO terms detected by at least two experimental approaches. We found that Tacrolimus, a calcineurin inhibitor via the immunophilin FKBP1A with properties similar to Cyclosporine, interacted with multiple genes and hubs in the Reactome mRNA Splicing pathway (Figure 7C). By assessing the KEGG drug interactome, we identified Zotatifin, a selective EIF4A inhibitor as an additional repurposed candidate drug targeting the Spliceosome. When evaluating the Reactome interferon signaling pathway and the late phase of HIV life cycle term, we detected Nintedanib, a triple angiokinase inhibitor used in the treatment of pulmonary fibrosis, as a candidate drug interacting with multiple genes and hubs of these pathways (*35*). To further assess the interaction network between the candidate drugs and their known biological targets, we intersected OmniPath’s protein-protein interaction database with the drugs known biological targets according to DrugBank (Figure 7D). Using genes detected by at least two assays as “proteins of interest”, we constructed the interaction network for Tacrolimus and Zotatifin for the mRNA splicing pathway and for Nintedanib for the interferon signaling pathway. Since the ‘Deceased’ patients exhibited an over-activation of these pathways, drugs interfering with the excessively activated pathways provide promising candidates for further study.

## DISCUSSION

We applied a systems biology approach to study severely ill COVID-19 patients at hospital admission. We hypothesized that a careful characterization of key immune effector cells using single cell transcriptomics and epigenomics would allow identifying host response pathways that lead to recovery or death. We identified monocytes as key effector cells of COVID-19 innate immunity and showed that detrimental host responses are epigenetically poised in the early stages of clinical deterioration. The importance of epigenetic changes of host cells for the viral life cycle had already been suggested by the finding of histone and chromatin modification during SARS-CoV-2 infection (*36*). Here, we expanded the epigenetics aspect of COVID-19 pathogenesis by applying an ***in-naturae*** approach where critically ill patients guided discovery of beneficial and detrimental host response pathways. Combining single cell transcriptomics with the epigenetics analysis, we identified dysregulated responses that constitute promising tractable pathways for pharmacological intervention to reduce mortality of hospitalized COVID-19 patients.

Our study design focused on comparing severely ill, hospitalized patients who survived to those who died. Thus, distinguishing itself from previous studies comparing severe COVID-19 patients to controls or mild patients (*37–41*); or studies comparing COVID-19 infected patients to other viral diseases (*41–43*). With our study design we were able to confirm well recognized COVID-19 severity characteristics such as type I IFN dysregulation (*44–46*); or the systemic upregulation of S100A8/A9 genes in monocytes (*38*). Furthermore, our results showed a strong correlation of PBMC composition with disease progression. Critically ill patients with poor prognosis showed a significant reduction of T cells and a significant increase of monocytes, consistent with previously reported findings in patients suffering from severe COVID-19 (*39, 47, 48*). The changes in the proportion of T cells and monocytes we observed were consistent with a declining role of innate immunity (monocytes) and a more prominent role of T cell immunity at the advanced stage of COVID-19 disease. Intriguingly, in monocytes and T cells, we detected a transcriptional signature at hospital admission which strongly correlated with disease evolution. At hospital admission, naive and central memory CD4+ T cells of patients with poor prognosis strong expression of genes enriched in “Apoptosis”, “Response to oxygen levels”, “Proteostasis” and “ER stress response” terms which are associated with early stages of cell death (*49, 50*). In monocytes of these patients, we observed a signature of reduced expression of genes involved in “Mitochondrial ATP Synthase” (mainly genes of the complex V), “MHC Complex” and “Ribosome and Translation Initiation” terms. Mitochondrial ATP activity is associated with cell cycle regulation and a decrease of ATP level is associated with cell proliferation (*51*). Overall, the concordance between the transcriptional signatures at an earlier stage of the disease and the changes of PBMC composition as the disease evolved reinforced the importance of early care treatment decisions to increase COVID-19 patient survival.

The extent of transcriptional variations detected between “Deceased” and “Alive” COVID-19 patients at hospital admission, and the continued transcriptional dysregulation with worsening disease at follow-up, supports the pivotal role of monocytes in COVID-19 severity and disease prognosis, as seen by others (*52*). The key role of monocytes for COVID-19 pathogenesis was also demonstrated by our finding that two of the five GWAS loci for respiratory failure co-localized with chromatin having increased accessibility in monocytes from patients who succumbed to the disease (*33*). We also showed that monocyte DNA methylation changes linked with disease severity are associated with genes in the extracellular matrix (ECM) and proteolysis pathways. The implication of the EMC in disease severity could highlight the damage to the structure and function of SARS-CoV-2 infected tissues (*53*), and the alteration of proteolysis pathways is consistent with the role of host proteolytic enzymes in viral replication and assembly (*54*). Strikingly, by combining monocyte transcriptomics with chromatin accessibility and DNA methylation, we identified among patients with equivalent clinical severity at admission, activated pathways that tagged those destined to succumb to COVID-19. Most prominent were pathways related to RNA splicing, metabolism and transport, chromatin architecture, and cellular response to interferon. Importantly, while the pathways were identified by a multi-omics approach in CD14+ monocytes, the corresponding transcriptomic changes could be tracked to B, T, and NK cells, indicating that their contribution is not limited to monocytes.

The functional pathways identified by the combined transcriptomics and epigenetics approach in our study had been previously implicated in COVID-9 pathogenesis. However, our study design pinpointed the differential activity of the pathways at the junction when potentially life-saving clinical treatment decisions for critically ill patients need to be made: when they are admitted to the intensive care unit. The involvement of interferon pathways in the COVID-19 pathogenesis has been well established (*44–46*). However, while we observed an increased IFN response in deceased patients, we cannot ascribe these abnormal responses as causal to the prognosis; alternatively, they may reflect a host compensatory mechanism (e.g. to uncontrolled viral replication). In contrast, RNA splicing by host cells is a key step of viral replication. SARS-CoV-2 encompasses 16 non-structural proteins (NSP1–NSP16) that are required for the virus to hijack the host transcriptional machinery. Viral NSP16 binds to the U1 and U2 spliceosome complex to negatively regulate host mRNA maturation (*55*). Additionally, *in vitro* challenge of Vero E6 cells with SARS-CoV-2 showed increased phosphorylation proteins involved in RNA splicing related pathways, positioning splicing as a critical mechanism of COVID-19 pathogenesis (*36*). The increased splicing activity observed for deceased patients might benefit viral replication and impair host immune responses. For instance, splicing disruption was shown to impair IFN responses to SARS-CoV-2 via intron retention (*55*). We found that genes encoding proteins involved in RNA transport and SRP-dependent protein targeting to the membrane were downregulated and less accessible in patients that would succumb to COVID-19. This may reflect the binding of viral NSP8 and NSP9 to the 7SL RNA component of the signal recognition particle (SRP) which interferes with protein trafficking (*55*).

Our study design further enabled the identification of candidate drugs to reduce mortality among critically ill COVID-19 patients. Using drug-protein and protein-protein interaction databases, we identified three drugs (Tacrolimus, Zotatifin, and Nintedanib) which target pathways that are differentially activated at hospital admission of critically ill patients and that discriminate between eventual survivors and decedents. Tacrolimus and Zotatifin may exert a dual role by interacting with the spliceosome and with NSPs from SARS-CoV-2 (*16, 56*). Although Tacrolimus is known as a calcineurin inhibitor, it also interacts with U2 RNA components of the spliceosome and was shown to bind to NSP1, a protein that SARS-CoV-2 uses to disrupt protein translation in infected cells (*56*). Additional support for Tacrolimus as a candidate drug are the observation that Cyclosporin (a molecule with similar properties to Tacrolimus) reduced mortality in hospitalized patients, and the increased survival from COVID-19 in liver transplant recipients treated with tacrolimus (*11, 57, 58*). Zotatifin, a member of the class of plant-derived cyclopenta[*b*]benzofurans compounds named rocaglates, inhibits specifically the RNA helicase eIF4A that is part of the EJC/TREX (exon junction and mRNA transport) complex of the spliceosome. Moreover, Zotatifin was shown in a network encompassing NSP13 and to interact with NSP9 (*16, 59*). The mechanism of action proposed for Zotatifin is via the reduction of viral infectivity and viral protein biogenesis and it is under clinical trial to treat COVID-19 (NCT04632381). Similarly to Zotatifin, the rocaglate CR-1-31-B was shown to potently inhibit SARS-CoV-2 replication at low nanomolar doses *in vitro* and *ex vivo* (*60*). Both Tacrolimus and Zotatifin were among drugs identified by *in vitro* and *in silico* models as candidate drugs to treat COVID-19 (*16, 17, 56*). Nintedanib is a drug used to treat pulmonary fibrosis. In a small clinical trial, nintedanib did not show significant improvement in the survival of severe pneumonia induced by COVID-19 (*61*). However, nintedanib shortened the mechanical ventilation time compared to the placebo group (*61*). Given that severely ill COVID-19 patients may require prolonged respiratory support, nintedanib could be considered as an adjunct therapy with other drugs. In our study, we confirmed the potential of Tacrolimus, Zotatifin, and Nintedanib to treat critically ill COVID-19 patients at hospital admission providing required pre-clinical data to support the testing of these drugs in controlled clinical studies.

## METHODS

### Patients

Subjects enrolled in the present study were individuals qPCR diagnosed with SARS-CoV-2 who were admitted to the McGill University Health Center (MUHC; Montreal, Quebec, Canada) and recruited to the Canadian Treatments for COVID-19 (CATCO) clinical trial (https://clinicaltrials.gov/ct2/show/NCT04330690). CATCO is a randomized, open-label, controlled trial and represents the Canadian arm of the WHO SOLIDARITY trial. Patient clinical progression was evaluated using the WHO Clinical Progression Scale, which represents a minimal common outcome measure set for COVID-19 clinical research (*32*). In brief, it provides a measure of illness severity from not infected (ordinal score of 0) to deceased (ordinal score of 10) (*32*). At the time of recruitment, patients in our cohort were randomized to receive either lopinavir/ritonavir (kaletra), or standard-of-care (Table 1). During the conduct of this study, the standard-of-care at our institution consisted solely of supplemental oxygen as required, antithrombotic prophylaxis, and supportive care in the intensive care unit when needed. Based on the outcome of hospitalization, patients were classified as “Deceased” if they died during hospitalization and “Alive” if they were discharged and considered recovered. At admission, all patients were severely ill (OS of 7). To follow disease evolution, disease severity during hospitalization was classified in 3 categories according to the WHO ordinal score: moderate (OS of 4 and 5); severe (OS of 6 or 7) and critical (OS of 8 or 9). Blood samples were collected through standard venipuncture in standard EDTA blood collection tubes and PBMC were obtained through Ficoll density centrifugation with SepMate tubes (Stemcell Technologies) and kept frozen in 10% DMSO + 90% FBS until analysis. The study was approved by the MUHC Research Ethics Board protocol 10-256.

### scRNA library preparation

Single cell PBMC suspensions were loaded on a Next GEM Chip G (PN-1000120) together with Next GEM Single Cell 3 GEM Kit v3.1 (PN-1000121) and single cells were captured on a 10X Genomics Chromium controller. cDNAs were generated following the 10X Genomics protocol. cDNAs were size selected using SPRIselect beads from Beckman Coulter (B23318), and their quality was checked with a Bioanalyzer High Sensitivity DNA Kit from Agilent (5067-4626). One quarter of the total cDNA was used to generate libraries using Chromium Next GEM Single Cell Library Kit (PN-1000121) and barcoded using the Single Index Kit Set A (PN-111213) following the 10X protocol. Libraries were size selected using SPRIselect beads and their quality was checked with the Bioanalyzer High Sensitivity DNA Kit. Libraries size were centered at 450 bp, and paired-end sequenced on NovaSeq 6000 S4 chips.

### ATAC-seq and WGBS library preparation from CD14+ monocytes

CD14+ monocytes were isolated from PBMCs using StemCell EasySep™ Human CD14 Positive Selection Kit II (17858). Viability was assessed with trypan blue staining, and only samples with viability > 85% were used. We then assessed chromatin accessibility using the ATAC-seq method employing 50,000 CD14+ monocytes (*62, 63*). Briefly, CD14+ monocytes were permeabilized with lysis buffer containing 0.05% IGEPAL and subsequently obtained nuclei were incubated with Tn5 transposase from Illumina (20034198) for 30 minutes at 37° C. Transposed DNAs were purified using Qiagen MinElute PCR Purification Kit and the ATAC-seq libraries were amplified for a total of 8-12 PCR cycles using NEBNext® High-Fidelity 2X PCR Master Mix (M0541) and the Nextera XT Index Kit v2 from Illumina (FC-131-2001, FC-131-2004). Final libraries were purified using sparQ PureMag Beads from QuantaBio (95196-060), and their quality was checked with High Sensitivity DNA Kit from Agilent (5067-4626). Libraries fragment sizes were mostly distributed between 200 bp and 1000 bp. Finally, libraries were 150 bp paired-end sequenced on a NovaSeq 6000 S4 chip.

Genomic DNA was extracted from CD14+ monocytes employing the Qiagen blood & tissue kit (69504). WGBS libraries were prepared using the NxSeq AmpFREE low DNA Library Kit. In summary genomic DNA was sheared using a Covaris E220 instrument. Post shearing cleaning-up was made using AMPure XP beads (0.8x) followed by an End-repair and an A-tailing to allow for adaptor ligation. After ligation, clean up is made using the AMPure XP beads (1x) before proceeding to the size selection (AMPure 0.75x). Another bead clean up (1X AMPure) is realized to remove primer dimers. Bisulfite conversion was made using the EZ-96 DNA Methylation-Gold^TM^ MagPrep from Zymo Research. The Bisulfite-Converted sample libraries were amplified using LM-PCR followed by a post-amplification clean up (AMPure 1x). Fifteen WGBS libraries were 150 bp paired-end sequenced on a NovaSeq 6000 S4 chip.

### Single-cell RNA-seq clustering analysis

Single-cell RNA-seq sequence reads quality was assessed with BVAtools (https://bitbucket.org/mugqic/bvatools/src/master/). Cell Ranger v3.0.1 was used for mapping the reads to the hg38 human reference genome assembly, filtering, and counting barcodes and UMIs, resulting in a list of UMI counts for every gene in every cell. To increase the robustness of our data analysis, we also integrated in the analysis 4 external healthy donor PBMC controls (3 males and 1 female) available from 10X Genomics (3 PBMC CITE-seq and 1 PBMC scRNA-seq). Data from 2 other healthy male donors generated on-site, where also included (*64*). Based on the barcode and UMI counts, we extracted the summary statistics for the number of genes expressed per cell and for the number of UMIs per cell and applied our cell quality and doublet filtering pipeline (*65*). Cells with >20% mitochondrial genes were excluded. Low-quality cells and doublets were filtered out by excluding cells falling outside the [-1 SD;+2.5 SD] interval for UMI and gene count distributions. Events co-expressing either of the cell specific markers TRBC1, LILRA4, CD14 and CD79A were considered doublets and excluded. The DoubletFinder package was used to identify and filter out the remaining cell doublets (*66*). Once dead cells and doublets had been removed, the COVID-19 patients and control samples were analyzed jointly with the Seurat v4 R package and cell clustering was performed by the uniform manifold approximation and projection dimension reduction method using the first 25 PCs with the most variable genes, but excluding mitochondrial and ribosomal protein genes (*67, 68*). This analysis identified 4 major populations divided into 22 distinct sub-populations, based on the marker genes for each cluster determined by the MAST approach and validated by TotalSeqB surface protein tagging by CITE-seq controls (*69*).

### Single-cell RNA-seq cellular composition analysis

To estimate the cellular composition, the PBMC lineage proportions were estimated for the 4 major populations and the 22 sub-populations. The number of cells composing each lineage inside a particular sample was normalized by the overall number of cells in that sample. The distribution of the sample specific lineage proportion was contrasted overall between Patients and Healthy Controls, at admission between patients retrospectively classified as “Deceased” and “Alive”, and at later time points between patients operationally defined as “Moderately” and “Critically” ill. Pairwise t-test comparisons for each contrast in each lineage was used to estimate the impact of COVID-19 on the cellular composition. FDR-adjusted p-values (q-value) were used to assess the significance of these comparisons.

### Single-cell RNA-seq differential gene expression (DGE) analysis

Gene expression changes across the two comparisons “Deceased vs Alive” at day 0 and “Critically vs Moderate” ill at follow-up (days 5 and 15) was evaluated in each lineage using the MAST approach adjusting for Age and Sex covariates (*69*). Adjusted p-values were estimated using the Benjamini-Hochberg (BH) procedure. Every significant differentially expressed gene (adj-p-value ≤0.05) was extracted as down- or up-regulated based on Log2 Fold Change value and used for downstream analysis. All differential expression results are provided in the Suppl. Table1.

### Single-cell RNA-seq pathway enrichment analysis

Enrichment analyses for GO biological processes were performed separately on up- and down-regulated DGE genes and for each cell sub-population using the enrichGO function of the clusterProfiler R package v3.13 (*70*). Enriched results were filtered to keep only GO terms for Biological Processes at level 5. Hierarchical level classification was obtained based on the directed acyclic graph (DAG) defined by the Gene Ontology Consortium provided through the GO.db R Package version v3.13. Additionally, only enrichment results showing a BH adjusted p-value ≤ 1e-5 supported by at least 5 differentially expressed genes were considered. Enrichment heatmaps were generated based on these selected results using the pheatmap R package v1.0.12. Lineage enrichment clustering was generated using the complete-linkage clustering method integrated in the pheatmap function. All level 5 enrichment results are provided in the Suppl. Table 2 (*71*).

### Single-cell RNA-seq Module Score analysis

Genes contributing to the enrichment of a GO category were extracted to form a transcriptional active module corresponding to that category. Module scores were estimated with the Seurat’s AddModuleScore function, using default settings (*72*). The Module score is determined for each cell by computing the mean transcriptional activity of the genes composing the module minus a mean expression extracted from a set of control genes. Control genes were randomly sampled from bins defined using the observed level of expression for the genes in the module. Module genes composition is available in Suppl. Table 7.

### ATAC-seq data analyses

The fifteen ATAC-seq libraries were checked for reads quality and low-quality reads were removed and adaptor sequences trimmed with Cutadapt v1.18 (*73*). Reads were then aligned to the human genome (hg38) using BWA v0.7.17 default parameters (*74*). PICARD v2.18.9 was used to mark duplicates and assess the fragment length distribution (*75*). Next, samtools v1.9 was used to remove reads aligned to mitochondria or alternative contigs (*76*). AlignmentSieve from DeepTools v3.5.0 was used to filter for unique paired reads (with --samFlagExclude 1804), to remove ENCODE hg38 blacklisted regions and to select fragments between 40 and 2000bp in length (*77, 78*). Library quality metrics were summarized with MultiQC v1.8 (*79*). The average fragment size per library was 219bp (SD 16.3bp) with a mean depth of 89.3 (SD 32.8) million fragments. All libraries presented a periodic nucleosome pattern and passed quality control. Chromatin accessibility was determined using MACS2 v2.2.6 callpeak with *q* value < 0.05 in BAMPE mode (*80*). We merged overlapping narrow peaks detected in at least two samples with bedtools v2.26 (*81*). The intersected peak set consisted of 13,892 genomic regions with an average width of 978bp. These peaks were selected for the count-based quantification of accessible chromatin. For each library, featureCounts v1.6.3 was used to count the number of unique fragments overlapping the targeted regions and determine the fraction of reads in peaks (FRIP) (*82*). We used filterByExpr from edgeR v3.30.3 to select 13,398 peaks with more than ten fragments in 80% of the 15 libraries. The filtered quantification matrix was normalized with the TMM method implemented on calcNormFactors (*83*). Counts were then scaled and normalized to log2 count per million (CPM) using limma-voom v3.44.3 (*84, 85*). The differential accessible chromatin (DAC) analysis for “Deceased vs Alive” at admission and at follow-up was performed with lmFit in limma (*84*). The linear model was adjusted on the covariates age, sex and FRIP. The log odds of differential accessibility was computed with eBayes (*84*). We estimated Storey’s false discovery rate (FDR) with qvalue v2.20.0 package (*86*). Peaks with FDR < 0.05 and absolute log2 fold change (log2FC) > 0.5 were considered significant DACs. To test if DAC regions were enriched in hospitalized GWAS loci, we used overlapPermTest from regionR v1.20.1 (*87*).

### ATAC-seq motif enrichment, gene ontology and pathway analysis

To test for enrichment of TF binding motifs in DAC regions, we used the findMotifsGenome function implemented in HOMER v4.11 (*88*). DAC regions in “Deceased vs Alive” at admision were compared to 50,000 random regions matched by DAC peak mean width and versus non-significant regions (FDR > 50%). We considered a motif to be significantly enriched if it presented FDR < 1% versus random and FDR < 10% versus non-significant regions in CD14+ monocytes. TFs were grouped by family and binding motif similarities after manual curation.

For GO and pathway analyses we assigned genes to peaks using two approaches. First, peaks were annotated to the genes with a transcription starting site (TSS) located within 5kb from the peak boundary using ChIPSeeker v1.24 (*89*). Next, we intersected our peak set with the GeneHancer v5 (2019) dataset using mergeByOverlaps from IRange package v2.22.2 (*90, 91*). By combining the TSS and GeneHancer approaches 76.7% of the 13,398 tested peaks were assigned to at least one gene. Enrichment analyses for KEGG, Reactome and GO biological processes were performed with clusterProfiler v3.16.0 and reactomePA v3.12 (*70, 92*). For the comparison of Open and Closed DAC regions at admission, pathways and GO terms with FDR < 1% and at least 10 genes were considered significant. For the follow-up and stratified analyses by motif groups an FDR < 5% and at least five genes in a pathway were deemed significant.

### Data visualization

To produce genomic tracks for ATAC-seq libraries we used bamCoverage from DeepTools (*78*) with a scaling factor based on edgeR’s TMM normalization method (*78*). This approach takes into account only fragments in peaks while also normalizing for library size. Next, SparK v2.6.2 was used to calculate the average base-pair accessibility and standard deviation per group (*93*). We used removeBatchEffect from limma to calculate the residual log2 CPM after regressing out covariates included in the linear models (*84*). Volcano plots were produced with ScatterView from MAGeCKFlute v1.8.0 (*94*). Raincloud, ggpubr and ggplot2 were used to produce the remaining ATAC-seq plots (*95, 96*).

### DNA methylation Calling and SNP Filtering

DNA methylation calling was performed with the GenPipes (v3.1.5-beta) “Methyl Seq’’ pipeline (*97*). In summary, raw reads were trimmed for quality (quality score ≥30) using trimmomatic (v0.36) and to remove sequencing adaptors (*73*), and then aligned to the human genome hg38 reference genome using Dynamic Read Analysis for Genomics (DRAGEN^TM^) epigenome pipeline (v3.4.5) which utilizes Illumina’s DRAGEN^TM^ Bio-It Platform with the improved and highly optimized mapping algorithms. All samples had a mean genome coverage of at least 18× (Suppl. Table 8). Duplicate reads were then removed using Picard (v2.9.0) [https://broadinstitute.github.io/picard/] and methylation calls were extracted using the methylation caller included as part of Bismark (v. 0.18.1) (*98*). CpGs were filtered to remove any position which overlap with a SNP loci, to reduce potential bias introduced by genetic variations. SNP loci were extracted from whole genome sequencing (WGS) libraries from the same patients. GenPipes (v3.1.6-beta) “DNA Seq’’ pipeline was used to analyze WGS data (*97*). Raw reads were trimmed for quality (quality score ≥ 30) using skewer 0.2.2 (PMID: 24925680), and then aligned to the human genome hg38 reference genome using BWA 0.7.17 with standard parameters. Alignment refinement was done using GATK (v3.8) and include marking duplicate, indel realignment and base recalibration process. The Haplotype_caller function from GATK (v3.8) was used to call variants (*99*). Every SNP found in at least one sample was used to define the set of SNP loci. On average, 860,127 (3.05%) CpGs were removed from each sample (Suppl. Table 8).

### Detection of differentially methylated CpG loci (DML) and differentially methylated regions (DMRs)

CpG sites with a minimum coverage of two in at least four samples were selected for differential analyses. Methylation raw data were smoothed using a 500-bp smoothing window as a parameter of the BSmooth function implemented in the bsseq Bioconductor package (v. 1.20.0) (*100*). Smoothed methylation levels of the CpG sites located on chromosome 21 of every sample were used to perform a principal component analysis to identify confounding factors. Differential methylation was detected using the R Bioconductor package DSS (v2.32.0) adjusting for patient age and sex (*101*). The model fitting was done by the DSS function DMLfit.multiFactor. Differentially methylated CpG sites (DML) were extracted using the DSS function DMLtest.multiFactor. Finally, DMLs with p-value < 0.05 were used to search differentially methylated regions (DMR) using the DSS function callDMR. The direction of the methylation changes (hyper- or hypo-methylation) was defined based on the sum of the test statistics of all DMLs within a DMR as provided by the areaStat value in the results of the callDMR function.

### Methylation motif enrichment, gene ontology and pathway analysis

The analysis of the enrichment of TF motifs in DMRs applied the same procedure as for the looking of TF enrichment in DAC ATAC-seq data. Only DMRs regions in the “Deceased vs Alive” comparison at admission were used to look for TF motifs enrichment. A motif was considered significantly enriched if the FDR was under 1% when compared to random regions.

To detect GO and pathway enrichments, we categorized DMRs into 3 groups based on their genomic location: Gene-body, Promoter and Distal DMRs. Gene-body DMRs are characterized by DMRs containing at least 10 CpGs located inside a gene. Promoter DMRs correspond to DMRs located in between −5kb and the +1kb from a gene transcriptional start site. Due to a high number of DMR located in promoter regions only the top 90% most differently methylated Promoter DMRs was conserved for the analysis. Finally, Distal DMRs were selected by overlapping Promoter DMRs with the GeneHancer v5 (2019) dataset using mergeByOverlaps from IRanges v2.22.2 package, and annotated based on the associated gene (*90*). For each of the 3 DMR categories, hyper- and hypo-methylated DMRs were analyzed separately. GO Biological Process and KEGG enrichments were detected using the clusterProfiler v3.16.0 package and Reactome enrichment using the reactomePA v1.32.0 package from R. Enrichments were considered significant at FDR <0.05 and if the enrichment is supported by at least 5 different genes and DMRs (*70, 92*).

### Data integration and identification of drug candidates

We performed data integration at the pathway and GO term levels. Pathways and GO terms detected by more than one assay with at least 10 genes and FDR < 5% (scRNA and ATAC) or 5 genes and FDR < 10% (DNA methylation) were considered for downstream drug discovery. Due to the hierarchical nature of GO terms and Reactome datasets we selected one term or pathway per hierarchical branch to reduce redundancies for plotting only (full datasets are provided in Suppl. Table 9).

To identify drugs interacting with genes and hubs in Reactome pathways we used ReactomeFIViz to access the DrugCentral database in Cytoscape (*102–104*). The DrugCentral database was created by collecting extensive drug information, including structure, active ingredients, mechanism of action and target interactions for more than 2000 FDA approved drugs. The KEGG database provides a section incorporating drug-pathway interactions, which was accessed to select candidate drugs for the Spliceosome pathway (*105*). KEGG drugs were curated according to their metabolism, transport, substrates, inhibitors and inducers to be assigned to a given pathway. To build a protein network around the targets for candidate drugs we used OmniPathR protein-protein interaction network and the DBparser packages to access the DrugBank dataset (*106, 107*). We started by extracting the proteins targeted by the candidate drugs in DrugBank. Next, we used genes detected by at least two assays to identify the interaction network starting from the drug target, passing through intermediary proteins, and ending in the “protein of interest”. The networks highlight the shortest interaction path between the drug target(s) to the proteins encoded by genes identified in our study. For Zotatifin and Tacrolimus, proteins of interest were encoded by genes part of the mRNA splicing pathway and for Nintedanib, genes in the Interferon Signaling pathway.

### Data availability

The scRNA-seq datasets for the hospitalized COVID-19 patients are available on the European Genome-Phenome Archive, accession number EGAS00001005468. The CITE-seq datasets from healthy controls were retrieved from 10X Genomics. The other healthy controls scRNA-seq datasets were from previously published studies (*108*). The ATAC-seq and WGBS datasets are available through the International Human Epigenome Consortium (IHEC) data portal (https://epigenomesportal.ca).

## Data Availability

The scRNA-seq datasets for the hospitalized COVID-19 patients are available on the European Genome-Phenome Archive, accession number EGAS00001005468. The CITE-seq datasets from healthy controls were retrieved from 10X Genomics. The other healthy controls scRNA-seq datasets were from previously published studies (L&eacutevy et al, JCI 2021). The ATAC-seq and WGBS datasets are available through the International Human Epigenome Consortium (IHEC) data portal (https://epigenomesportal.ca).

## ACKNOWLEDGMENTS

The authors are highly grateful to the study participants for their contribution as well as acknowledge support received by individuals working at the McGill Genome Center and the McGill Epigenomics Mapping Center (EMC). This work was supported by a Canada Institute of Health Research (CIHR) program grant (CEE-151618) for the McGill EMC, which is part of the Canadian Epigenetics, Environment and Health Research Consortium (CEEHRC) Network and a CIHR Project Grant to DL (#168959). DL was also supported by an FRQ-S, Chercheur-Boursier Junior 1 Award and the Canada Foundation for Innovation John R. Evans Leaders Fund. Work in the lab of ES was supported by a Foundation grant from CIHR (FDN-143332) and a Canada Foundation for Innovation John R. Evans Leaders Fund award. GB is supported by a Canada Research Chair Tier 1 award, a FRQ-S, Distinguished Research Scholar award and the Canadian Center for Computational Genomics (C3G) is supported by a Genome Canada Genome Technology Platform grant. MPC was supported by a CIHR grant during the conduct of the study. The work was also supported by an award from the McGill Interdisciplinary Initiative in Infection and Immunity (MI4) to MPC, MB, GB, and ES. This research was enabled in part by support provided by Calcul Quebec and Compute Canada (www.computecanada.ca).

