## Supplemental Figures for "A system biology approach identifies candidate drugs to reduce mortality in severely ill COVID-19 patients"

### SUPPLEMENTAL MATERIAL

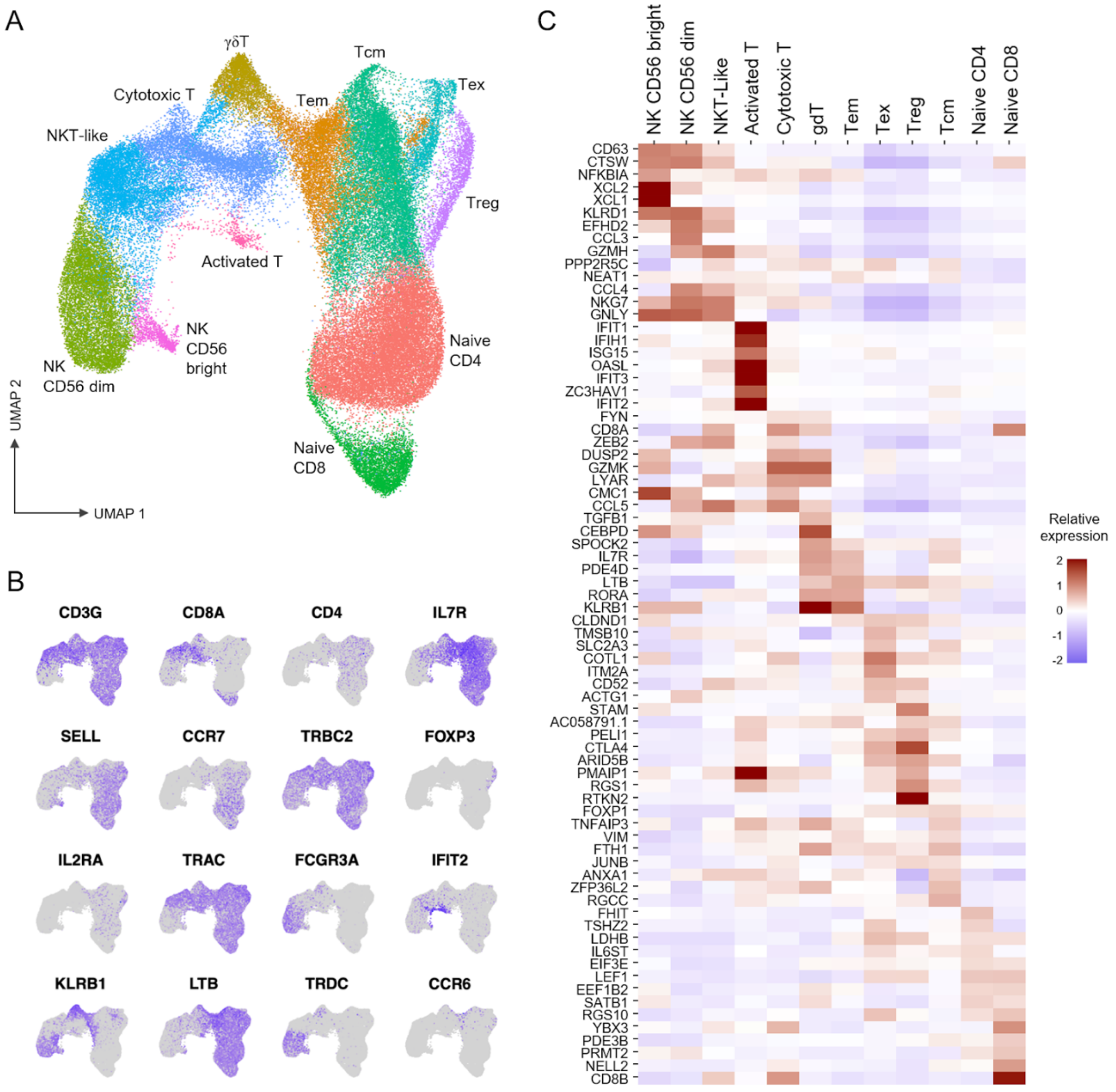


**Suppl. Figure 1. Sub-population composition of T and NK cells from COVID-19 patients.**

**(A)** UMAP visualization for the subclustering of T and NK cells selected PBMC of 7 COVID-19 patients (18 samples obtained at days 0, 5 and 15 post admission) and 6 healthy controls. Twelve major sub-populations were identified. **(B)** UMAP visualization of sixteen major T cell marker genes used to validate sub-populations annotation. **(C)** Heatmap representation of the normalized expression in each individual T-cell subpopulation for the top 74 genes identified as sub-population specific markers using the Seurat 'FindAllMarkers' function.

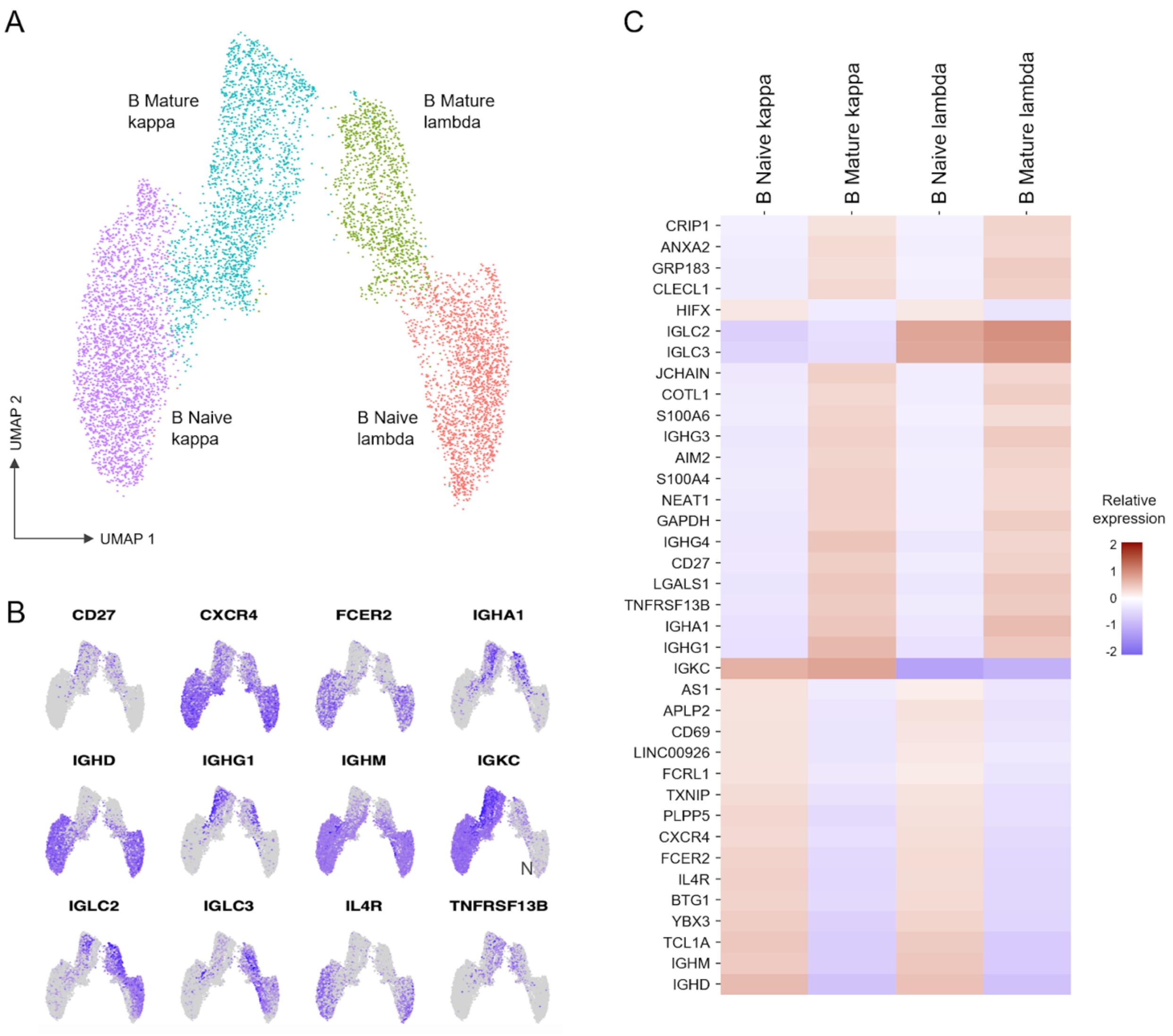


**Suppl. Figure 2. Sub-population composition of B cells from COVID-19 patients.**

**(A)** UMAP visualization of B cell sub-populations selected from 7 COVID-19 patients (18 samples obtained at days 0, 5 and 15 of admission) and 6 healthy controls. Four major sub-populations are identified. **(B)** UMAP visualization of twelve major B-cell marker genes used to validate sub-population annotation. **(C)** Heatmap representation of the normalized expression in each individual B-cell subpopulation for the top 37 genes identified as sub-population specific markers using the Seurat 'FindAllMarkers' function.

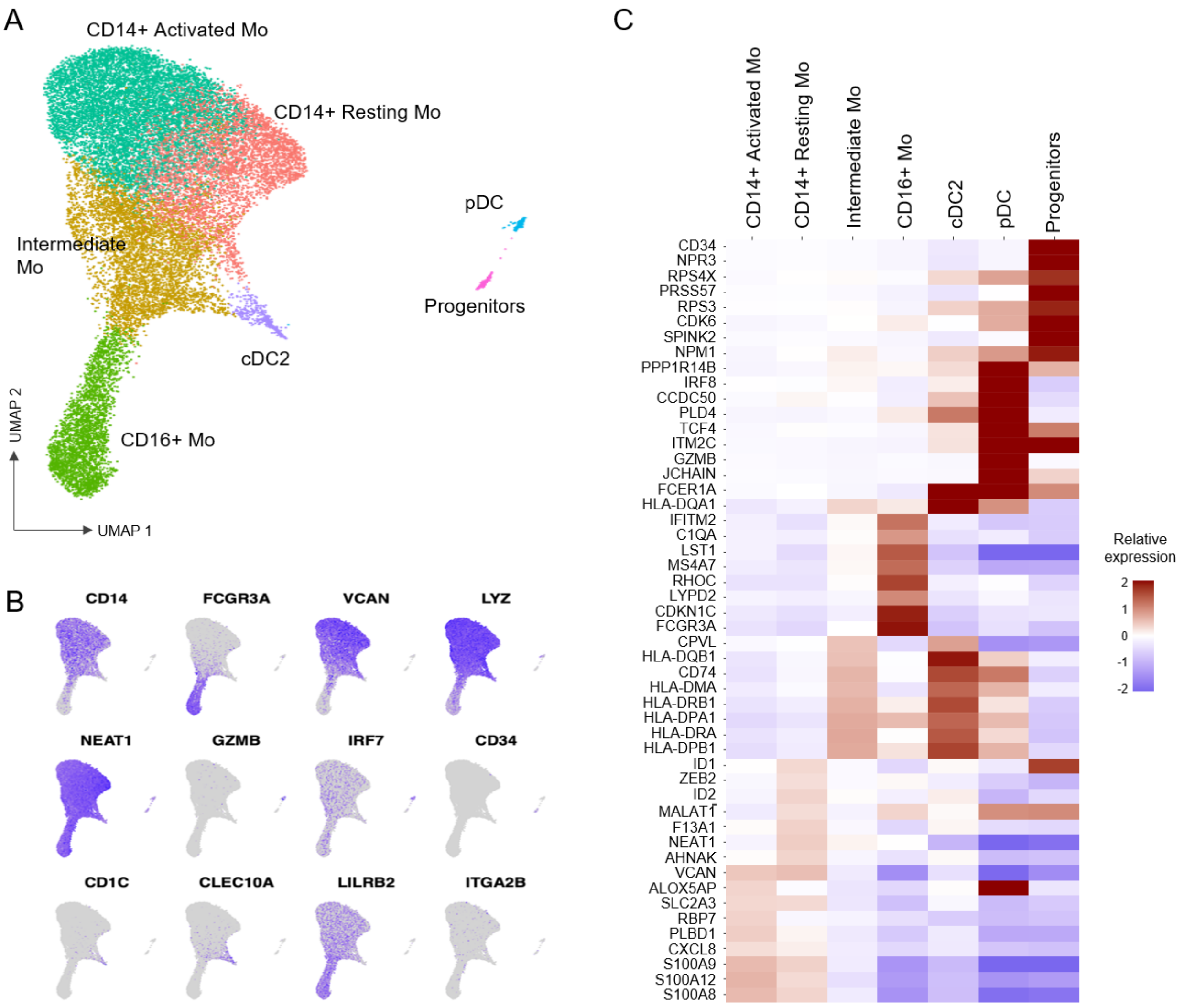


**Suppl. Figure 3. Sub-population composition of Myeloid cells from COVID-19 patients.**

**(A)** UMAP visualization of Myeloid cells subclustering selected from PBMC of 7 COVID-19 patients (18 samples obtained at days 0, 5 and 15 of admission) and 6 healthy controls. Seven major sub-populations were identified. **(B)** UMAP visualization of twelve marker genes used to validate sub-population annotation. **(C)** Heatmap representation of the normalized expression in each Myeloid cell subpopulation for the top 50 genes identified as sub-population specific markers using the Seurat 'FindAllMarkers' function.

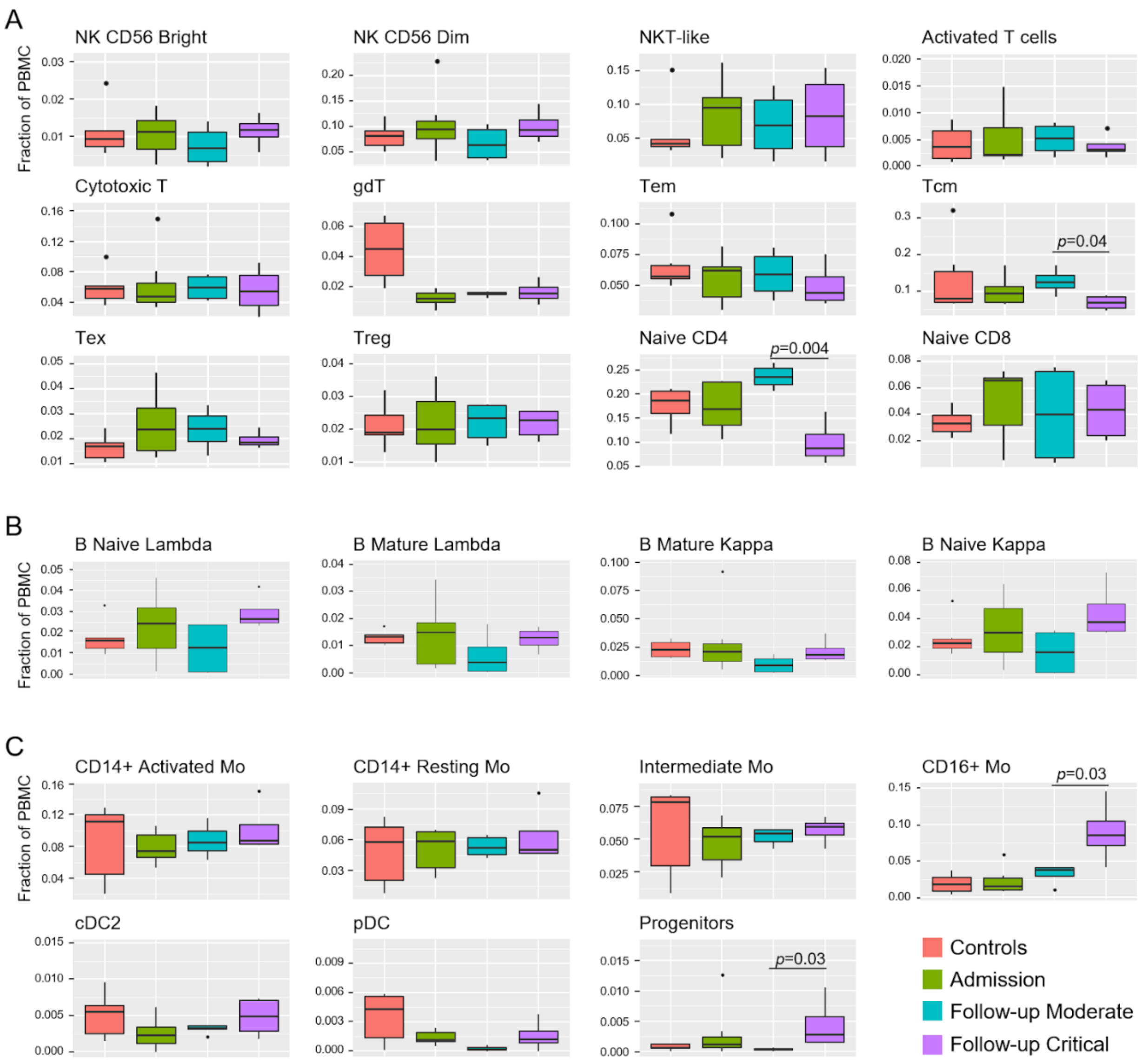


**Suppl. Figure 4. PBMC sub-population cell proportions**

Box and whisker plots for the proportion of cells for each subpopulation of T cells **(A)**, B cells **(B)** and Myeloid cells **(C)** for samples in each clinical group; expressed as % of total PBMC. Lineage proportions at days 5 and 15 of COVID-19 patients classified based on the disease OS and grouped as Moderate(cyan) or Critical(purple). Corresponding values from healthy controls (red) and from all COVID-19 patients at admission (green) are provided as reference groups. Q-values for pairwise t-test between Critical and Moderate comparisons are provided when significant.

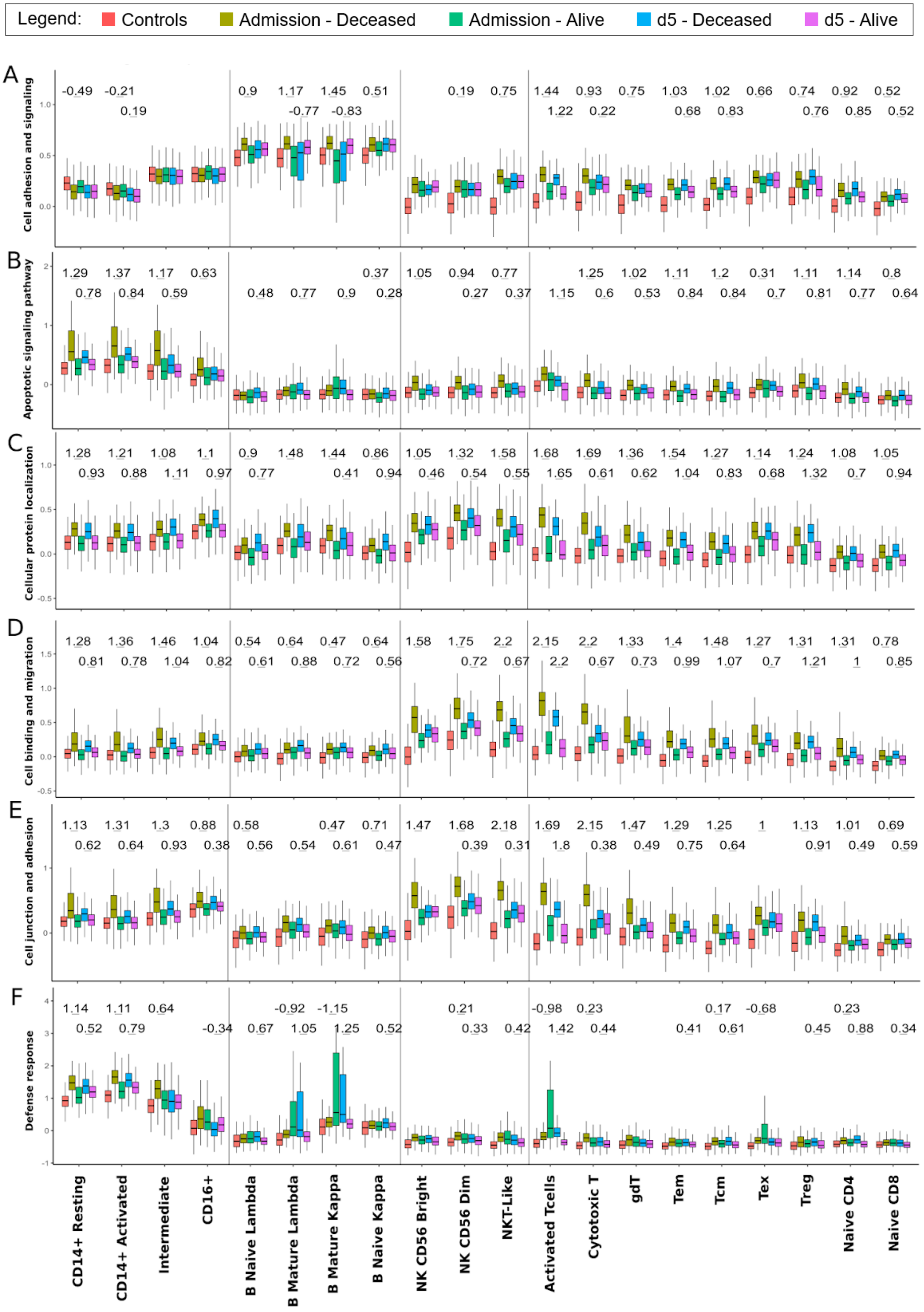

Legend: Controls Admission - Deceased Admission - Alive d5 - Deceased d5 - Alive
A
Cell adhesion and signaling
-0.49 -0.21 0.9 1.17 1.45 0.51 0.19 0.75 1.44 0.93 0.75 1.03 1.02 0.66 0.74 0.92 0.52
0.19 -0.77 -0.83 1.22 0.22 0.68 0.83 0.76 0.85 0.52
B
Apoptotic signaling pathway
1.29 1.37 1.17 0.63 0.37 1.05 0.94 0.77 1.25 1.02 1.11 1.2 0.31 1.11 1.14 0.8
0.78 0.84 0.59 0.48 0.77 0.9 0.28 0.27 0.37 1.15 0.6 0.53 0.84 0.84 0.7 0.81 0.77 0.64
C
Cellular protein localization
1.28 1.21 1.08 1.1 0.9 1.48 1.44 0.86 1.05 1.32 1.58 1.68 1.69 1.36 1.54 1.27 1.14 1.24 1.08 1.05
0.93 0.88 1.11 0.97 0.77 0.41 0.94 0.46 0.54 0.55 1.65 0.61 0.62 1.04 0.83 0.68 1.32 0.7 0.94
D
Cell binding and migration
1.28 1.36 1.46 1.04 0.54 0.64 0.47 0.64 1.58 1.75 2.2 2.15 2.2 1.33 1.4 1.48 1.27 1.31 1.31 0.78
0.81 0.78 1.04 0.82 0.61 0.88 0.72 0.56 0.72 0.67 2.2 0.67 0.73 0.99 1.07 0.7 1.21 1 0.85
E
Cell junction and adhesion
1.13 1.31 1.3 0.88 0.58 0.47 0.71 1.47 1.68 2.18 1.69 2.15 1.47 1.29 1.25 1 1.13 1.01 0.69
0.62 0.64 0.93 0.38 0.56 0.54 0.61 0.47 0.39 0.31 1.8 0.38 0.49 0.75 0.64 0.91 0.49 0.59
F
Defense response
1.14 1.11 0.64 -0.92 -1.15 0.21 -0.98 0.23 0.17 -0.68 0.23
0.52 0.79 -0.34 0.67 1.05 1.25 0.52 0.33 0.42 1.42 0.44 0.41 0.61 0.45 0.88 0.34
CD14+ Resting
CD14+ Activated
Intermediate
CD16+
B Naive Lambda
B Mature Lambda
B Mature Kappa
B Naive Kappa
NK CD56 Bright
NK CD56 Dim
NKT-Like
Activated Tcells
Cytotoxic T
gdT
Tem
Tcm
Tex
Treg
Naive CD4
Naive CD8

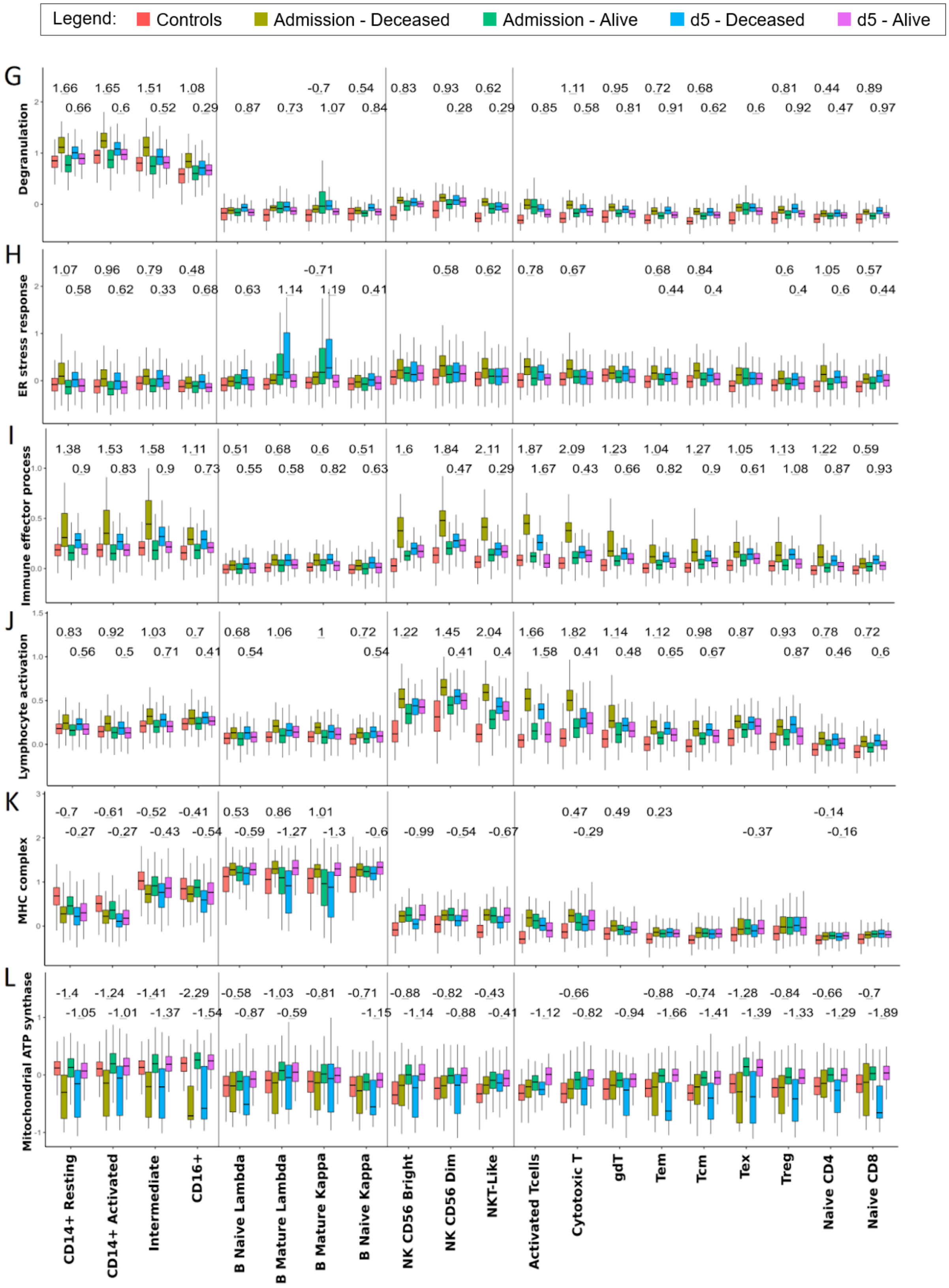

Legend: Controls Admission - Deceased Admission - Alive d5 - Deceased d5 - Alive
G
Degranulation
1.66 1.65 1.51 1.08 -0.7 0.54 0.83 0.93 0.62 1.11 0.95 0.72 0.68 0.81 0.44 0.89
0.66 0.6 0.52 0.29 0.87 0.73 1.07 0.84 0.28 0.29 0.85 0.58 0.81 0.91 0.62 0.6 0.92 0.47 0.97
H
ER stress response
1.07 0.96 0.79 0.48 -0.71 0.58 0.62 0.78 0.67 0.68 0.84 0.6 1.05 0.57
0.58 0.62 0.33 0.68 0.63 1.14 1.19 0.41 0.44 0.4 0.4 0.6 0.44
I
Immune effector process
1.38 1.53 1.58 1.11 0.51 0.68 0.6 0.51 1.6 1.84 2.11 1.87 2.09 1.23 1.04 1.27 1.05 1.13 1.22 0.59
0.9 0.83 0.9 0.73 0.55 0.58 0.82 0.63 0.47 0.29 1.67 0.43 0.66 0.82 0.9 0.61 1.08 0.87 0.93
J
Lymphocyte activation
0.83 0.92 1.03 0.7 0.68 1.06 1 0.72 1.22 1.45 2.04 1.66 1.82 1.14 1.12 0.98 0.87 0.93 0.78 0.72
0.56 0.5 0.71 0.41 0.54 0.54 0.41 0.4 1.58 0.41 0.48 0.65 0.67 0.87 0.46 0.6
K
MHC complex
-0.7 -0.61 -0.52 -0.41 0.53 0.86 1.01 0.47 0.49 0.23 -0.14
-0.27 -0.27 -0.43 -0.54 -0.59 -1.27 -1.3 -0.6 -0.99 -0.54 -0.67 -0.29 -0.37 -0.16
L
Mitochondrial ATP synthase
-1.4 -1.24 -1.41 -2.29 -0.58 -1.03 -0.81 -0.71 -0.88 -0.82 -0.43 -0.66 -0.88 -0.74 -1.28 -0.84 -0.66 -0.7
-1.05 -1.01 -1.37 -1.54 -0.87 -0.59 -1.15 -1.14 -0.88 -0.41 -1.12 -0.82 -0.94 -1.66 -1.41 -1.39 -1.33 -1.29 -1.89
CD14+ Resting
CD14+ Activated
Intermediate
CD16+
B Naive Lambda
B Mature Lambda
B Mature Kappa
B Naive Kappa
NK CD56 Bright
NK CD56 Dim
NKT-Like
Activated Tcells
Cytotoxic T
gdT
Tem
Tcm
Tex
Treg
Naive CD4
Naive CD8

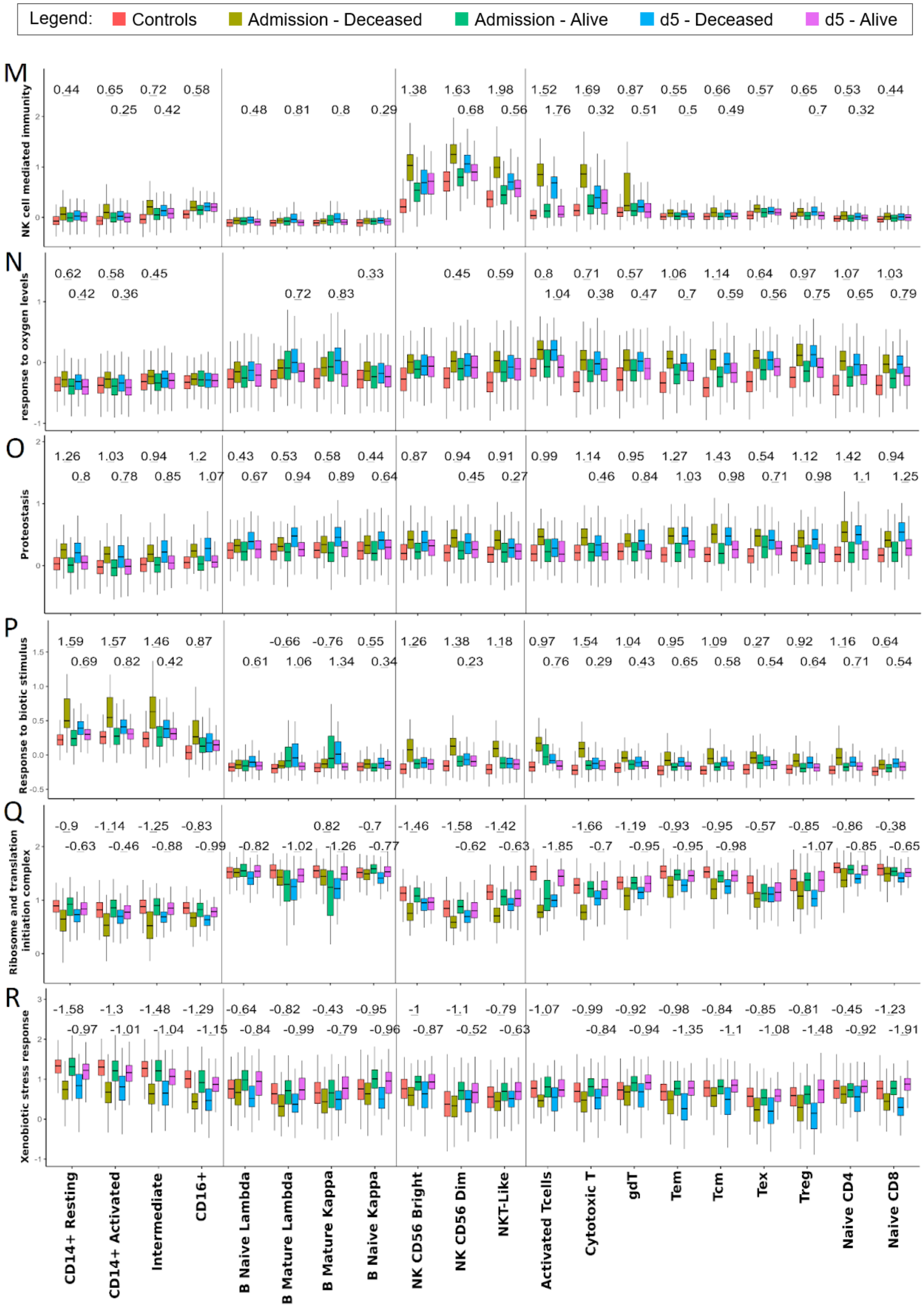
Legend: Controls Admission - Deceased Admission - Alive d5 - Deceased d5 - Alive
M
NK cell mediated immunity
N
response to oxygen levels
O
Proteostasis
P
Response to biotic stimulus
Q
Ribosome and translation initiation complex
R
Xenobiotic stress response
CD14+ Resting
CD14+ Activated
Intermediate
CD16+
B Naive Lambda
B Mature Lambda
B Mature Kappa
B Naive Kappa
NK CD56 Bright
NK CD56 Dim
NKT-Like
Activated Tcells
Cytotoxic T
gdT
Tem
Tcm
Tex
Treg
Naive CD4
Naive CD8

**Suppl. Figure 5. Evolution of the transcriptional expression for enriched GO-terms.**

Differentially expressed genes contributing to GO term enrichment (indicated on the left axis and displayed in Figure 4A) have been used to generate a Module Score representing the overall transcriptional expression level for the specific category. Corresponding cell populations are listed at the bottom of the graph. Module scores have been computed for the Day 0 and Day 5 time-points for each cell population and based on "Deceased" and "Alive" COVID-19 patient outcomes separately. Control samples from healthy subjects are included as reference. Sample groups are coloured according to the key on top of the graph. The significance of the gene expression differences between "Deceased vs Alive" COVID-19 patients was tested by comparing the corresponding Module Score distribution in every group of cells using a wilcoxon rank-test approach (q-value <= 0.05). To provide a sample-size free evaluation of the difference in expression, Cohen's d effect size estimation is shown above each significant variation observed.

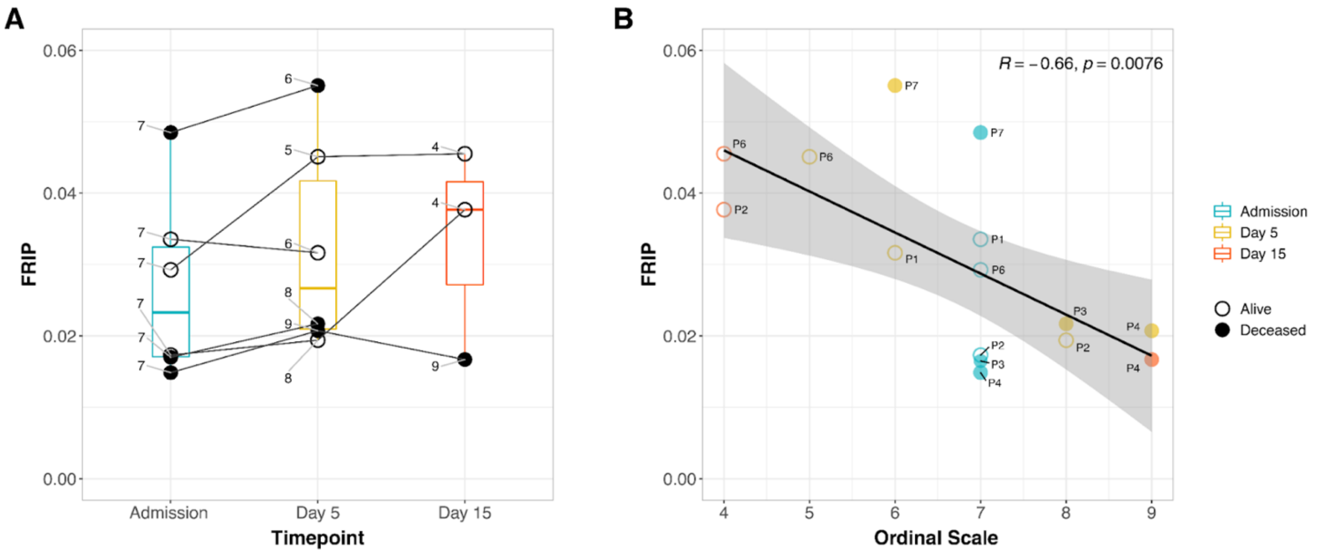


**Suppl. Figure 6. COVID-19 WHO clinical ordinal scale correlates with chromatin stability of CD14+ monocytes.**

**(A)** The fraction of reads in peaks (FRIP) for CD14+ monocytes, a surrogate for chromatin stability, was plotted against the date of PBMC collection. The WHO ordinal scale for COVID-19 is shown for all patients at each timepoint. **(B)** Correlation of the FRIP with WHO clinical scale score for COVID-19. Samples are labeled according to the donor ID, day of collection and patient outcome.

#### 716 More close peaks in deceased at Admission

| Name | MOTIF | # of DAC with Motif | % of DAC with Motif | # random peaks with Motif | % of random peaks with Motif | # peaks at FDR > 50% with Motif | % peaks at FDR > 50% with Motif | p-value | FDR q-value |
|---|---|---|---|---|---|---|---|---|---|
| **Group 1 - NFY** | | | | | | | | | |
| NFY | | 282 | 49.3% | 12676 | 27.4% | 1356 | 34.9% | 1.0E-28 | < 5.0E-05 |
| **Group 2 - Sp1-like/KLF** | | | | | | | | | |
| Sp1 | | 297 | 51.9% | 14991 | 32.4% | 1592 | 40.9% | 1.0E-21 | < 5.0E-05 |
| KLF3 | | 308 | 53.9% | 16188 | 34.9% | 1642 | 42.2% | 1.0E-19 | < 5.0E-05 |
| KLF1 | | 404 | 70.6% | 24860 | 53.7% | 2318 | 59.6% | 1.0E-16 | < 5.0E-05 |
| Klf9 | | 238 | 41.6% | 13083 | 28.2% | 1256 | 32.3% | 1.0E-11 | < 5.0E-05 |
| Sp5 | | 409 | 71.5% | 27005 | 58.3% | 2488 | 64.0% | 1.0E-10 | < 5.0E-05 |
| Klf4 | | 189 | 33.0% | 10073 | 21.7% | 968 | 24.9% | 1.0E-09 | < 5.0E-05 |
| KLF5 | | 419 | 73.3% | 29349 | 63.4% | 2612 | 67.2% | 1.0E-06 | < 5.0E-05 |
| KLF6 | | 389 | 68.0% | 26803 | 57.9% | 2389 | 61.4% | 1.0E-06 | < 5.0E-05 |
| Egr2 | | 136 | 23.8% | 7583 | 16.4% | 671 | 17.3% | 1.0E-05 | < 5.0E-05 |
| KLF10 | | 193 | 33.7% | 12843 | 27.7% | 1100 | 28.3% | 1.0E-03 | 5.6E-03 |
| Maz | | 412 | 72.0% | 30442 | 65.7% | 2566 | 66.0% | 1.0E-03 | 4.5E-03 |
| **Group 3 - E2F** | | | | | | | | | |
| E2F4 | | 273 | 47.7% | 17136 | 37.0% | 1629 | 41.9% | 1.0E-07 | < 5.0E-05 |
| E2F7 | | 101 | 17.7% | 5881 | 12.7% | 556 | 14.3% | 1.0E-03 | 2.8E-03 |
| E2F6 | | 274 | 47.9% | 18963 | 40.9% | 1642 | 42.2% | 1.0E-03 | 3.0E-03 |
| E2F1 | | 173 | 30.2% | 11181 | 24.1% | 996 | 25.6% | 1.0E-03 | 3.3E-03 |
| E2F3 | | 298 | 52.1% | 20918 | 45.2% | 1818 | 46.8% | 1.0E-03 | 3.3E-03 |
| **Group 4 - MYB** | | | | | | | | | |
| MYB | | 369 | 64.5% | 23351 | 50.4% | 2064 | 53.1% | 1.0E-11 | < 5.0E-05 |
| AMYB | | 302 | 52.8% | 19292 | 41.6% | 1815 | 46.7% | 1.0E-07 | < 5.0E-05 |
| **Group 5 - CRE** | | | | | | | | | |
| CRE | | 114 | 19.9% | 5163 | 11.1% | 599 | 15.4% | 1.0E-09 | < 5.0E-05 |
| **Group 6 - HOX** | | | | | | | | | |
| Hoxa9 | | 362 | 63.3% | 24219 | 52.3% | 2058 | 52.9% | 1.0E-07 | < 5.0E-05 |
| Hoxd10 | | 190 | 33.2% | 11890 | 25.7% | 1069 | 27.5% | 1.0E-04 | 3.0E-04 |
| **Group 7 - PBX** | | | | | | | | | |
| Pknox1 | | 75 | 13.1% | 3567 | 7.7% | 308 | 7.9% | 1.0E-05 | 1.0E-04 |
| Pbx3 | | 66 | 11.5% | 3201 | 6.9% | 263 | 6.8% | 1.0E-04 | 4.0E-04 |
| PBX1 | | 30 | 5.2% | 1072 | 2.3% | 84 | 2.2% | 1.0E-04 | 4.0E-04 |
| MafA | | 165 | 28.9% | 10200 | 22.0% | 896 | 23.0% | 1.0E-04 | 7.0E-04 |
| **Group 8 - ZNF189** | | | | | | | | | |
| ZNF189 | | 203 | 35.5% | 12778 | 27.6% | 1056 | 27.2% | 1.0E-04 | 3.0E-04 |
| **Group 9 - LIM homeobox** | | | | | | | | | |
| En1 | | 271 | 47.4% | 18020 | 38.9% | 1600 | 41.1% | 1.0E-04 | 3.0E-04 |
| Isl1 | | 280 | 49.0% | 18856 | 40.7% | 1596 | 41.0% | 1.0E-04 | 4.0E-04 |
| Lhx3 | | 230 | 40.2% | 15145 | 32.7% | 1241 | 31.9% | 1.0E-04 | 8.0E-04 |
| **Group 10 - Oct4:Sox17** | | | | | | | | | |
| Oct4:Sox17 | | 33 | 5.8% | 1222 | 2.6% | 65 | 1.7% | 1.0E-04 | 3.0E-04 |
| **Group 11 - FOX** | | | | | | | | | |
| Foxh1 | | 115 | 20.1% | 6695 | 14.5% | 556 | 14.3% | 1.0E-03 | 1.2E-03 |
| Foxo1 | | 288 | 50.4% | 19992 | 43.2% | 1734 | 44.6% | 1.0E-03 | 2.3E-03 |
| **Group 12 - GATA** | | | | | | | | | |
| GATA | | 32 | 5.6% | 1394 | 3.0% | 120 | 3.1% | 1.0E-03 | 4.6E-03 |
| **Group 13 - TBX** | | | | | | | | | |
| Tbx20 | | 56 | 9.8% | 2936 | 6.3% | 258 | 6.6% | 1.0E-03 | 5.8E-03 |
| **Group 14 - NKX** | | | | | | | | | |
| Arnt:Ahr | | 194 | 33.9% | 12608 | 27.2% | 1154 | 29.7% | 1.0E-03 | 1.8E-03 |
| Nkx2.2 | | 336 | 58.7% | 24193 | 52.2% | 1946 | 50.0% | 1.0E-03 | 5.9E-03 |
| Nkx2.1 | | 404 | 70.6% | 29952 | 64.7% | 2395 | 61.6% | 1.0E-02 | 7.9E-03 |
| **Group 15 - ZNF669** | | | | | | | | | |
| ZNF669 | | 45 | 7.9% | 2290 | 4.9% | 170 | 4.4% | 1.0E-02 | 9.4E-03 |

#### 243 More open peaks in deceased at Admission

| Name | MOTIF | # of DAC with Motif | % of DAC with Motif | # random peaks with Motif | % of random peaks with Motif | # peaks at FDR > 50% with Motif | % peaks at FDR > 50% with Motif | p-value | FDR q-value |
|---|---|---|---|---|---|---|---|---|---|
| **Group 16 - IRF** | | | | | | | | | |
| PU.1:IRF8 | | 35 | 16.6% | 2597 | 5.2% | 346 | 9.8% | 1.0E-08 | < 5.0E-05 |
| IRF8 | | 43 | 20.4% | 4285 | 8.7% | 541 | 15.3% | 1.0E-06 | < 5.0E-05 |
| IRF2 | | 17 | 8.1% | 1279 | 2.6% | 193 | 5.5% | 1.0E-04 | 1.5E-03 |
| IRF4 | | 37 | 17.5% | 4711 | 9.5% | 451 | 12.8% | 1.0E-03 | 3.8E-03 |
| **Group 17 - CTCF** | | | | | | | | | |
| CTCF | | 30 | 14.2% | 3261 | 6.6% | 324 | 9.2% | 1.0E-04 | 1.8E-03 |
| BORIS | | 53 | 25.1% | 7884 | 15.9% | 603 | 17.1% | 1.0E-03 | 5.9E-03 |
| **Group 18 - MADS** | | | | | | | | | |
| CArG | | 29 | 13.7% | 3423 | 6.9% | 236 | 6.7% | 1.0E-03 | 5.2E-03 |

**Suppl Figure 7. Motif homology groping for the 46 transcription factors enriched in DAC regions of deceased patients at Admission.**

The 46 transcription factors significantly enriched in DAC regions of deceased patients at FDR $<1\%$ are shown with their respective motif binding sequences in each row. Transcription factors were grouped based on the sequence homology of their binding motif. The number and percentage of peaks with motifs corresponding to the heatmap in figure 4D is shown for DAC, random peaks, and peaks at FDR $> 50\%$ in the center columns. Uncorrected *p*-value and the FDR q-value for the contrast between DAC and random regions is given in the last two columns.

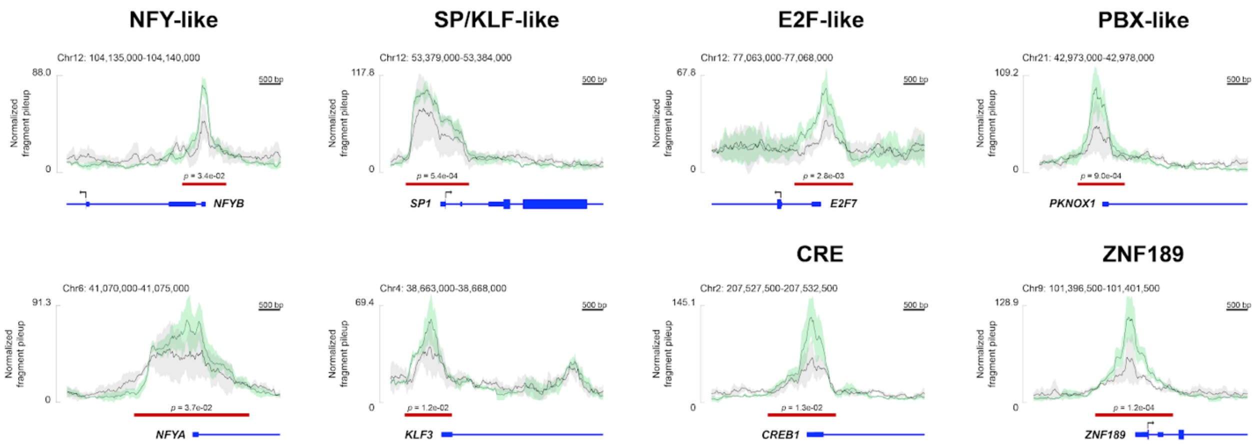


**Suppl Figure 8. Chromatin accessibility at promoters of transcription factors with binding motifs enriched in repressed DAC regions of deceased COVID-19 patients.**

The mean chromatin accessibility at admission for patients retrospectively classified as Deceased or Alive are shown as black and green lines, respectively. Shades indicate the standard deviation of the mean for each group. At the bottom, a red bar denotes the regions contrasted in the “Deceased vs Alive” comparison with its corresponding *p* value.

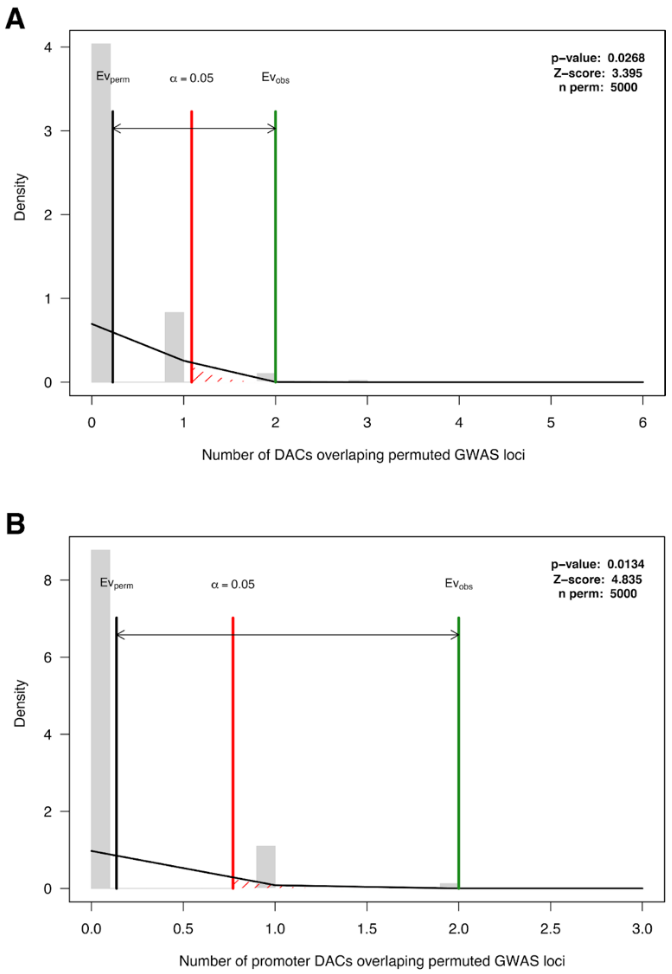


**Suppl Figure 9. Non-random overlap between DAC for "Deceased" patient monocytes at admission with hospitalized COVID-19 GWAS loci.**

The evaluation of a non-random overlap between DAC regions with five significant/suggestive GWAS loci for hospitalized COVDI-19 patients was performed using a permutation test. Genomic regions of equal length to the five GWAS loci were permuted across the human genome while the genomic position for all 959 "Deceased vs Alive" DAC regions for monocytes at admission **(A)**, and for the subset of 481 DAC located in promoter regions **(B)**. The bar plots summarize the number of permuted GWAS loci overlapping a DAC region in the x-axis with the grey bars representing the density plotted in the y-axis. The black vertical line ($Ev_{perm}$) indicates the average number of permuted GWAS loci encompassing a DAC; the red line represents the 95 percentile of the distribution; and the green line ($Ev_{obs}$) highlights the number of GWAS loci with a DAC in our study.

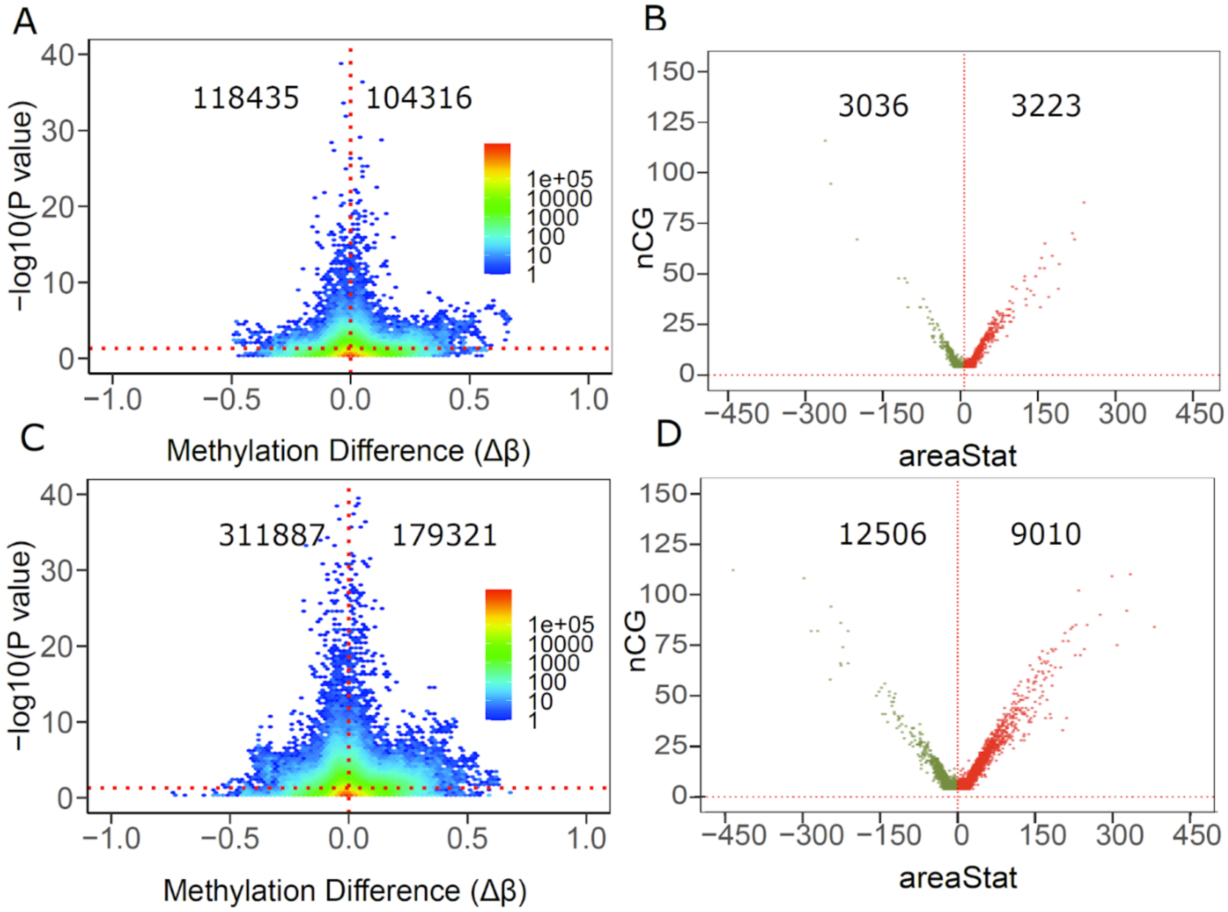


**Suppl Figure 10. Differential DNA methylation among COVID-19 patients.**

**(A)** A total of 222,751 DMLs were identified for the "Deceased vs Alive" patient comparison at admission. Hyper ($\Delta\beta > 0$) and hypomethylated ($\Delta\beta <= 0$) CpGs were plotted against their corresponding *p* values. The color gradient indicates the density of CpGs. **(B)** AreasStats of hyper (areaStat > 0) and hypomethylated (areaStat < 0) regions were plotted against the number of CpGs located in the DMR for "Deceased vs Alive" patients at admission (red indicates hypermethylated DMRs and olive green indicates hypomethylated DMRs). **(C)** A total of 491,208 DMLs were detected for the "Deceased vs Alive" patient groups at follow-up. Hyper and hypomethylated CpGs were plotted against their corresponding *p* values. **(D)** AreasStats of hyper (areaStat > 0) and hypomethylated (areaStat < 0) regions were plotted against the number of CpGs located in the DMR for "Deceased vs Alive" patients at follow-up.

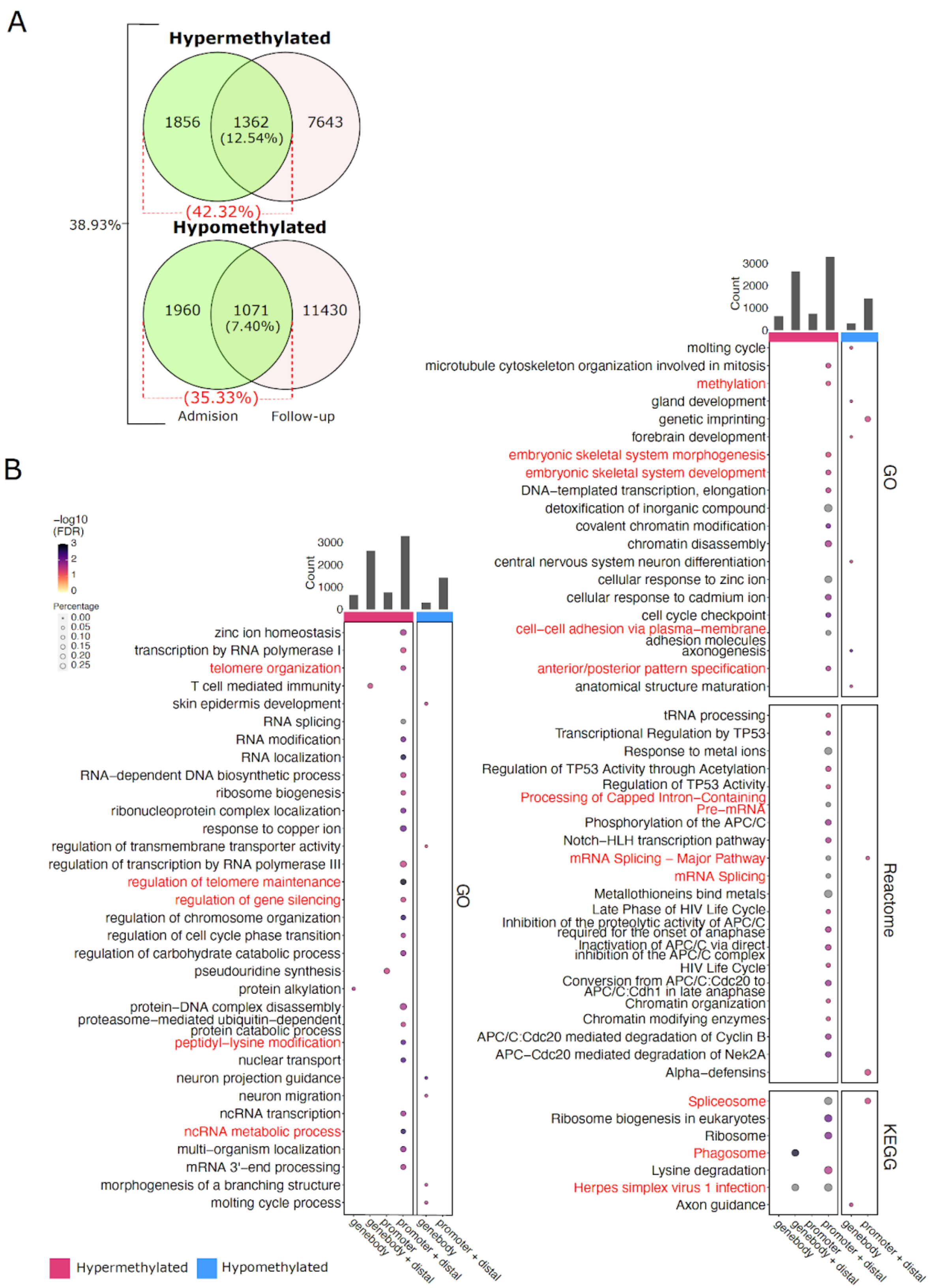

A
Hypermethylated
1856
1362
(12.54%)
7643
(42.32%)
38.93%
Hypomethylated
1960
1071
(7.40%)
11430
(35.33%)
Admision
Follow-up
B
−log10 (FDR)
Percentage
Count
zinc ion homeostasis
transcription by RNA polymerase I
telomere organization
T cell mediated immunity
skin epidermis development
RNA splicing
RNA modification
RNA localization
RNA−dependent DNA biosynthetic process
ribosome biogenesis
ribonucleoprotein complex localization
response to copper ion
regulation of transmembrane transporter activity
regulation of transcription by RNA polymerase III
regulation of telomere maintenance
regulation of gene silencing
regulation of chromosome organization
regulation of cell cycle phase transition
regulation of carbohydrate catabolic process
pseudouridine synthesis
protein alkylation
protein−DNA complex disassembly
proteasome−mediated ubiquitin−dependent protein catabolic process
peptidyl−lysine modification
nuclear transport
neuron projection guidance
neuron migration
ncRNA transcription
ncRNA metabolic process
multi−organism localization
mRNA 3'−end processing
morphogenesis of a branching structure
molting cycle process
GO
genebody
genebody + distal
promoter
promoter + distal
Hypermethylated
Hypomethylated
molting cycle
microtubule cytoskeleton organization involved in mitosis
methylation
gland development
genetic imprinting
forebrain development
embryonic skeletal system morphogenesis
embryonic skeletal system development
DNA−templated transcription, elongation
detoxification of inorganic compound
covalent chromatin modification
chromatin disassembly
central nervous system neuron differentiation
cellular response to zinc ion
cellular response to cadmium ion
cell cycle checkpoint
cell−cell adhesion via plasma−membrane adhesion molecules
axonogenesis
anterior/posterior pattern specification
anatomical structure maturation
tRNA processing
Transcriptional Regulation by TP53
Response to metal ions
Regulation of TP53 Activity through Acetylation
Regulation of TP53 Activity
Processing of Capped Intron−Containing Pre−mRNA
Phosphorylation of the APC/C
Notch−HLH transcription pathway
mRNA Splicing − Major Pathway
mRNA Splicing
Metallothioneins bind metals
Late Phase of HIV Life Cycle
Inhibition of the proteolytic activity of APC/C required for the onset of anaphase
Inactivation of APC/C via direct inhibition of the APC/C complex
HIV Life Cycle
Conversion from APC/C:Cdc20 to APC/C:Cdh1 in late anaphase
Chromatin organization
Chromatin modifying enzymes
APC/C:Cdc20 mediated degradation of Cyclin B
APC−Cdc20 mediated degradation of Nek2A
Alpha−defensins
Reactome
Spliceosome
Ribosome biogenesis in eukaryotes
Ribosome
Phagosome
Lysine degradation
Herpes simplex virus 1 infection
Axon guidance
KEGG

**Suppl Figure 11: Comparison of monocytes DNA methylation at admission and follow-up. (A)** Number and percentages of overlapping DMRs between monocytes of patients classified on the disease outcome ("Deceased vs Alive") at the time of admission and follow-up. Overall intersecting percentages for both hyper and hypomethylated DMRs are shown on the left. **(B)** Gene ontology and pathway enrichment analyses for DMRs at follow-up. Each bubble indicates an ontology or pathway for each of the three tested databases: KEGG and Reactome pathways, and Gene Ontology. The bubble size represents the percentage of genes with a corresponding significant DMR at FDR <5% and shades represent the -log10(FDR). The total number of peaks with assigned genes per group is shown in the bar plot on the right. Pathways also enriched at admission are colored in red.

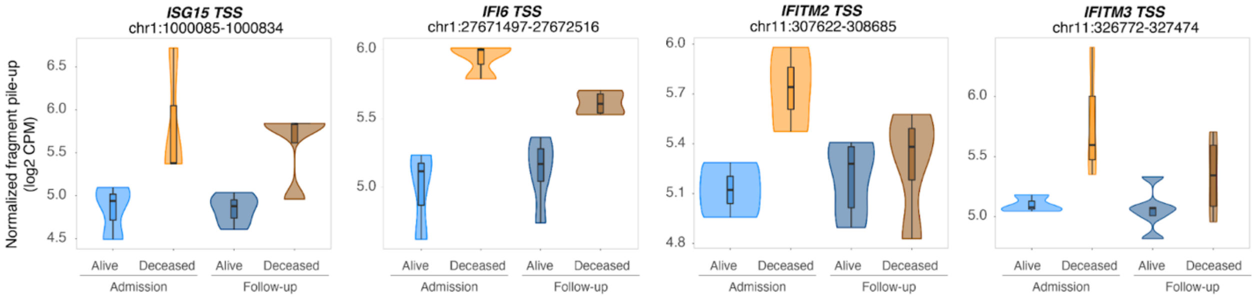


**Suppl. Figure 12: Genes in type-1 interferon signaling GO term show increased promoter accessibility.**

Box plots with normalized quantification of pileup fragments in peaks at promoter regions for "Deceased" and "Alive" are shown as log2 count per million (CPM) for patient groups at admission and follow-up.

#### SUPPLEMENTAL TABLES

**Suppl. Table S1:** Differential gene expression analysis for each immune cell subpopulation identified by scRNAseq.

**Suppl. Table S2:** Gene ontology and pathway enrichment analysis for differentially expressed genes per sub-population.

**Suppl. Table S3:** Differential accessible chromatin analysis in monocytes of hospitalized COVID-19 patients.

**Suppl. Table S4: Suppl. Table S4:** Gene ontology and pathway enrichment analysis for differentially accessible chromatin in monocytes.

**Suppl. Table S5:** List of differentially methylated regions in monocytes of hospitalized COVID-19 patients at the admission.

**Suppl. Table S6:** List of differentially methylated regions in monocytes of hospitalized COVID-19 patients at the follow-up.

**Suppl. Table S7:** List of genes included in the Modules Scores.

**Suppl. Table S8:** WGBS sequencing coverage and SNP removal statistics.

**Suppl. Table S9:** Intersection of gene ontology and pathway enrichment analyses.
